## Appendices for "Accuracy and clinical effectiveness of risk prediction tools for pressure injury occurrence: An umbrella review"

### S1 File: Appendices

#### Appendix 1: PRISMA-DTA Checklist

| Section/topic | # | PRISMA-DTA Checklist Item | Where reported (paragraph #) |
|---|---|---|---|
| **TITLE / ABSTRACT** | | | |
| Title | 1 | Identify the report as a systematic review (+/- meta-analysis) of ~~diagnostic test~~ accuracy ~~(DTA) studies~~ *reviews* | Title page and abstract |
| Abstract | 2 | Abstract: See PRISMA-DTA for abstracts. | Abstract |
| **INTRODUCTION** | | | |
| Rationale | 3 | Describe the rationale for the review in the context of what is already known. | Introduction (1-3) |
| Clinical role of index test | D1 | State the scientific and clinical background, including the intended use and clinical role of the index test, and if applicable, the rationale for minimally acceptable test accuracy (or minimum difference in accuracy for comparative design). | Introduction (1-3) |
| Objectives | 4 | Provide an explicit statement of question(s) being addressed in terms of participants, index test(s), and target condition(s). | Introduction (4) |
| **METHODS** | | | |
| Protocol and registration | 5 | Indicate if a review protocol exists, if and where it can be accessed (e.g., Web address), and, if available, provide registration information including registration number. | Methods: Protocol registration and reporting of findings |
| Eligibility criteria | 6 | Specify study characteristics (participants, setting, index test(s), reference standard(s), target condition(s), and study design) and report characteristics (e.g., years considered, language, publication status) used as criteria for eligibility, giving rationale. | Methods: Eligibility criteria for this umbrella review |
| Information sources | 7 | Describe all information sources (e.g., databases with dates of coverage, contact with study authors to identify additional studies) in the search and date last searched. | Methods: Literature search |
| Search | 8 | Present full search strategies for all electronic databases and other sources searched, including any limits used, such that they could be repeated. | Appendix 2 |
| Study selection | 9 | State the process for selecting studies (i.e., screening, eligibility, included in systematic review, and, if applicable, included in the meta-analysis). | Methods: Literature search, Eligibility criteria |
| Data collection process | 10 | Describe method of data extraction from reports (e.g., piloted forms, independently, in duplicate) and any processes for obtaining and confirming data from investigators. | Methods: Data extraction and quality assessment (1) |
| Definitions for data extraction | 11 | Provide definitions used in data extraction and classifications of target condition(s), index test(s), reference standard(s) and other characteristics (e.g. study design, clinical setting). | Methods: Data extraction and quality assessment (1); |

### PRISMA-DTA Checklist

| Section/topic | # | PRISMA-DTA Checklist Item | Where reported (paragraph #) |
|---|---|---|---|
| | | | Appendix 3 |
| Risk of bias and applicability | 12 | Describe methods used for assessing risk of bias in individual studies and concerns regarding the applicability to the review question. | Methods: Data extraction and quality assessment (2); Appendix 4 |
| Diagnostic accuracy measures | 13 | State the principal diagnostic accuracy measure(s) reported (e.g. sensitivity, specificity) and state the unit of assessment (e.g. per-patient, per-lesion). | Methods: Data extraction and quality assessment (1); Appendix 3 |
| Synthesis of results | 14 | Describe methods of handling data, combining results of studies and describing variability between studies. This could include, but is not limited to: a) handling of multiple definitions of target condition. b) handling of multiple thresholds of test positivity, c) handling multiple index test readers, d) handling of indeterminate test results, e) grouping and comparing tests, f) handling of different reference standards | Methods: Synthesis methods |
| Meta-analysis | D2 | Report the statistical methods used for meta-analyses, if performed. | -- |
| Additional analyses | 16 | Describe methods of additional analyses (e.g., sensitivity or subgroup analyses, meta-regression), if done, indicating which were pre-specified. | -- |
| **RESULTS** | | | |
| Study selection | 17 | Provide numbers of studies screened, assessed for eligibility, included in the review (and included in meta-analysis, if applicable) with reasons for exclusions at each stage, ideally with a flow diagram. | Figure 1; Appendix 5 Table A1 |
| Study characteristics | 18 | For each included study provide citations and present key characteristics including: a) participant characteristics (presentation, prior testing), b) clinical setting, c) study design, d) target condition definition, e) index test, f) reference standard, g) sample size, h) funding sources | Table 1; Table 2; Table 4; Appendix 5 Table A2 |
| Risk of bias and applicability | 19 | Present evaluation of risk of bias and concerns regarding applicability for each study. | Figure 2; Appendix 5 Table A3 |
| Results of individual ~~studies~~ *reviews* | 20 | For each analysis in each ~~study~~ *review* (e.g. unique combination of index test, reference standard, and positivity threshold) report ~~2x2 data (TP, FP, FN, TN) with~~ estimates of diagnostic accuracy and confidence intervals, ~~ideally with a forest or receiver operator characteristic (ROC) plot.~~ | Table 3; Appendix 5 Tables A4-6 |

| Section/topic | # | PRISMA-DTA Checklist Item | Where reported (paragraph #) |
|---|---|---|---|
| Synthesis of results | 21 | Describe test accuracy, including variability; if meta-analysis was done, include results and confidence intervals. | Results: Results from reviews evaluating the prognostic accuracy of risk prediction tools – Results from reviews evaluating the clinical effectiveness of risk prediction tools |
| Additional analysis | 23 | Give results of additional analyses, if done (e.g., sensitivity or subgroup analyses, meta-regression; analysis of index test: failure rates, proportion of inconclusive results, adverse events). | -- |
| **DISCUSSION** | | | |
| Summary of evidence | 24 | Summarize the main findings including the strength of evidence. | Discussion (1);<br>Discussion: Prognostic accuracy of risk prediction tools – Clinical effectiveness of risk prediction scales |
| Limitations | 25 | Discuss limitations from included studies (e.g. risk of bias and concerns regarding applicability) and from the review process (e.g. incomplete retrieval of identified research). | Discussion (1);<br>Discussion: Strengths and limitations |
| Conclusions | 26 | Provide a general interpretation of the results in the context of other evidence. Discuss implications for future research and clinical practice (e.g. the intended use and clinical role of the index test). | Conclusions |
| **FUNDING** | | | |
| Funding | 27 | For the systematic review, describe the sources of funding and other support and the role of the funders. | Funding |

*Adapted From:* McInnes MDF, Moher D, Thombs BD, McGrath TA, Bossuyt PM, The PRISMA-DTA Group (2018). Preferred Reporting Items for a Systematic Review and Meta-analysis of Diagnostic Test Accuracy Studies: The PRISMA-DTA Statement. JAMA. 2018 Jan 23;319(4):388-396. doi: 10.1001/jama.2017.19163.

#### Appendix 2: Description of search strategies

ORIGINAL SEARCH: JAN 2023

**Summary table of searches**

| Source | Results before deduplication | Results after deduplication |
|---|---|---|
| MEDLINE | 1643 | 574 |
| EMBASE | 2060 | 1920 |
| CINAHL | 3720 | 3007 |
| EPISTEMONIKOS | 1194 | 574 |
| GOOGLE SCHOLAR | 357 | 226 |
| **TOTAL** | **8974** | **6301** |

SEARCH UPDATE: JUNE 2024

**Searches run from 01/01/23 - 06/24**

**Summary table of searches**

| Source | Results before deduplication | Results after deduplication |
|---|---|---|
| MEDLINE | 229 | 186 |
| EMBASE | 330 | 175 |
| CINAHL | 480 | 383 |
| EPISTEMONIKOS | 232 | 118 |
| GOOGLE SCHOLAR | 80 | 36 |
| **TOTAL** | **1351** | **898** |

**Search approach and sources**

Search concepts:

- **i.** pressure injury terms
- **ii.** systematic review terms
- **iii.** prediction model terms

Pressure injury (PI) terms were used from previous PI topic reviews and were developed in consultation with the wider review team and customer.

Established systematic review methodological filters were used in OVID Embase and OVID MEDLINE combining the appropriate McMasters best balance reviews filters[i] combining the appropriate McMasters best balance systematic reviews filters[ii] with the appropriate CADTH systematic review filter[iii].

A number of existing methodological filters are available for prediction/prognostic model terms. The effect of using different combinations of these filters have been tested in order to ensure retrieval of relevant literature at a manageable volume. This testing has informed the choice of prognostic search filters used (Geersing)[iv] Haynes Best Balance[v] and Ingui Best Balance[vi].

Searches were run in OVID MEDLINE, OVID Embase and EBSCO CINAHL Plus using the search concepts, systematic review and prediction/prognostic filters listed above or adaptations of these filters. No publication date or language restrictions were applied.

Epistemonikos was also searched using PI terms and key prognostic terms limited by publication type systematic review or broad synthesis. The Epistemonikos interface does not support the same search functionality available in OVID or EBSCO (for example adjacency operators are not supported). The Information Specialist ran several separate shorter searches to accommodate for the limitations of the interface. No publication date or language restrictions were applied.

In addition, Google Scholar was searched to pick up any potentially relevant papers not indexed in the other databases. The Google Scholar interface has limited search functionality. The Information Specialist ran several separate shorter searches to accommodate for the limitations of the interface. Searches were limited to review publication types published in the last eleven years only for pragmatic reasons as Google Scholar has poor export functionality.

"Connected papers" was also considered for inclusion, however it is a one 'seed tool', i.e., searching for one paper generates one map of connected papers. The platform is also only freely accessible for

---

[i] Search strategies for EMBASE in Ovid Syntax, In Health Information Research Unit Hedges project.Ontario:HIRU;2022: Health Information Research Unit - HIRU ~ Search Strategies for EMBASE in Ovid Syntax (mcmaster.ca) Accessed 2022-09-29 and
Search strategies for MEDLINE in Ovid Syntax, In Health Information Research Unit Hedges project.Ontario:HIRU;2022: Health Information Research Unit - HIRU ~ Search Strategies for MEDLINE in Ovid Syntax and the PubMed translation (mcmaster.ca) Accessed 2022-09-29.

[ii] Search strategies for EMBASE in Ovid Syntax, In Health Information Research Unit Hedges project.Ontario:HIRU;2022: Health Information Research Unit - HIRU ~ Search Strategies for EMBASE in Ovid Syntax (mcmaster.ca) Accessed 2022-09-29 and
Search strategies for MEDLINE in Ovid Syntax, In Health Information Research Unit Hedges project.Ontario:HIRU;2022: Health Information Research Unit - HIRU ~ Search Strategies for MEDLINE in Ovid Syntax and the PubMed translation (mcmaster.ca) Accessed 2022-09-29.

[iii] SR / MA / HTA / ITC - MEDLINE, Embase, PsycInfo. In: CADTH Search Filters Database. Ottawa: CADTH; 2022: https://searchfilters.cadth.ca/link/33. Accessed 2022-09-29.

[iv] Geersing GJ, Bouwmeester W, Zuithoff P, et al. Search filters for finding prognostic and diagnostic prediction studies in Medline to enhance systematic reviews. *PLoS One* 2012;7(2):e32844. doi:10.1371/journal.pone.0032844 [published Online First: 2012/03/07]

[v] Wilczynski NL, Haynes RB. Optimal search strategies for detecting clinically sound prognostic studies EMBASE: and analytic survey. *J Am Med Inform Assoc* 2005;12(4):481-5. doi: 10.1197/jamia.M1752 [published Online First: 2005/04/02]

[vi] Ingui BJ, Rogers MA. Searching for clinical prediction rules in MEDLINE. *J Am Med Inform Assoc* 2001;8(4):391-7. doi: 10.1136/jamia.2001.0080391 [published Online First: 2001/06/22]

searching five ‘seed’ papers a month, which appears to be more of a limitation than would be beneficial for this set of reviews.

**MEDLINE ALL (OVID)**

**Date run: 31/01/23**

Database: Ovid MEDLINE(R) ALL <1946 to January 30, 2023>

Search Strategy:

--------------------------------------------------------------------------------

1 (decubit* or bedsore* or bed-sore* or pressure-ulcer* or pressure-wound*).tw. (17989)

2 ((pressure* or bed or bedbound or bed-bound or bedridden or bed-ridden or deep tissue* or deep-tissue) adj3 (wound* or ulcer* or sore* or injur* or lesion*)).tw. (22136)

3 exp pressure ulcer/ or pressure/ae (15198)

4 1 or 2 or 3 (33198)

5 ((supine or immobil*) adj3 (heal or healing or heals or healed or dress*)).tw. (220)

6 ((supine or immobil*) adj3 (wound* or ulcer* or sore* or injur* or lesion*)).tw. (876)

7 ((pressure or bedbound or bedridden or bed-bound or bed-ridden or deep tissue or deep-tissue) adj3 (heal or healing or heals or healed or dress*)).tw. (1863)

8 5 or 6 or 7 (2923)

9 4 or 8 (34931)

10 (systematic review or meta-analysis).pt. (299072)

11 review.pt. (3115171)

12 search:.tw. (617577)

13 meta-analys:.mp. (291117)

14 meta-analysis/ or systematic review/ or systematic reviews as topic/ or meta-analysis as topic/ or "meta analysis (topic)"/ or "systematic review (topic)"/ or exp technology assessment, biomedical/ or network meta-analysis/ (336517)

15 ((systematic* adj3 (review* or overview*)) or (methodologic* adj3 (review* or overview*))).ti,ab,kf. (303236)

16 ((quantitative adj3 (review* or overview* or synthes*)) or (research adj3 (integrati* or overview*))).ti,ab,kf. (15021)

17 ((integrative adj3 (review* or overview*)) or (collaborative adj3 (review* or overview*)) or (pool* adj3 analy*)).ti,ab,kf. (37395)

18 (data synthes* or data extraction* or data abstraction*).ti,ab,kf. (38583)

19 (handsearch* or hand search*).ti,ab,kf. (10921)

20 (mantel haenszel or peto or der simonian or dersimonian or fixed effect* or latin square*).ti,ab,kf. (34465)

21 (met analy* or metanaly* or technology assessment* or HTA or HTAs or technology overview* or technology appraisal*).ti,ab,kf. (11813)

22 (meta regression* or metaregression*).ti,ab,kf. (13870)

23 (meta-analy* or metaanaly* or systematic review* or biomedical technology assessment* or bio-medical technology assessment*).mp,hw. (446528)

24 (medline or cochrane or pubmed or medlars or embase or cinahl).ti,ab,hw. (325867)

25 (cochrane or (health adj2 technology assessment) or evidence report).jw. (21207)

26 (comparative adj3 (efficacy or effectiveness)).ti,ab,kf. (17070)

27 (outcomes research or relative effectiveness).ti,ab,kf. (11017)

28 ((indirect or indirect treatment or mixed-treatment or bayesian) adj3 comparison*).ti,ab,kf. (4214)

29 (multi* adj3 treatment adj3 comparison*).ti,ab,kf. (287)

30 (mixed adj3 treatment adj3 (meta-analy* or metaanaly*)).ti,ab,kf. (177)

31 umbrella review*.ti,ab,kf. (1305)

32 (multi* adj2 paramet* adj2 evidence adj2 synthesis).ti,ab,kf. (13)

33 (multiparamet* adj2 evidence adj2 synthesis).ti,ab,kf. (18)

34 (multi-paramet* adj2 evidence adj2 synthesis).ti,ab,kf. (11)

35 or/10-34 (3717455)

36 predict:.tw. or validat:.mp. or develop.tw. (3128744)

37 (stratification or ROC curve).ti,ab. or exp ROC curve/ or discriminat$.ti,ab. or c-statistic.ti,ab. or "Area under the curve".ti,ab. or AUC.ti,ab. or Calibration.ti,ab. or indices.ti,ab. or algorithm.ti,ab. or multivaria$.mp. (1567005)

38 Validat*.mp. or Predict$.ti. or Rule*.mp. or (Predict* and (Outcome* or Risk* or Model$)).mp. or ((History or Variable$ or Criteria or Scor$ or Characteristic$ or Finding$ or Factor$) and (Predict$ or Model$ or Decision$ or Identif$ or Prognos$)).mp. or (Decision$.mp. and ((Model$ or Clinical$).mp. or Logistic Models/)) or (Prognostic and (History or Variable$ or Criteria or Scor$ or Characteristic$ or Finding$ or Factor$ or Model$)).mp. [mp=title, book title, abstract, original title, name of substance word, subject heading word, floating sub-heading word, keyword heading word, organism supplementary concept word, protocol supplementary concept word, rare disease supplementary concept word, unique identifier, synonyms] (5961362)

39 36 or 37 (6673746)

40 39 or 38 (7368991)

41 9 and 35 and 40 (1643)

**MEDLINE ALL (OVID)**

**Date run: 20/06/24**

Database: Ovid MEDLINE(R) ALL <1946 to June 20, 2024>

Search Strategy:

--------------------------------------------------------------------------------

1 (decubit* or bedsore* or bed-sore* or pressure-ulcer* or pressure-wound*).tw.

2 ((pressure* or bed or bedbound or bed-bound or bedridden or bed-ridden or deep tissue* or deep-tissue) adj3 (wound* or ulcer* or sore* or injur* or lesion*)).tw.

3 exp pressure ulcer/ or pressure/ae

4 1 or 2 or 3

5 ((supine or immobil*) adj3 (heal or healing or heals or healed or dress*)).tw.

6 ((supine or immobil*) adj3 (wound* or ulcer* or sore* or injur* or lesion*)).tw.

7 ((pressure or bedbound or bedridden or bed-bound or bed-ridden or deep tissue or deep-tissue) adj3 (heal or healing or heals or healed or dress*)).tw.

8 5 or 6 or 7

9 4 or 8

10 (systematic review or meta-analysis).pt.

11 review.pt.

12 search:.tw.

13 meta-analys:.mp.

14 meta-analysis/ or systematic review/ or systematic reviews as topic/ or meta-analysis as topic/ or "meta analysis (topic)"/ or "systematic review (topic)"/ or exp technology assessment, biomedical/ or network meta-analysis/

15 ((systematic* adj3 (review* or overview*)) or (methodologic* adj3 (review* or overview*))).ti,ab,kf.

16 ((quantitative adj3 (review* or overview* or synthes*)) or (research adj3 (integrati* or overview*))).ti,ab,kf.

17 ((integrative adj3 (review* or overview*)) or (collaborative adj3 (review* or overview*)) or (pool* adj3 analy*)).ti,ab,kf.

18 (data synthes* or data extraction* or data abstraction*).ti,ab,kf.

19 (handsearch* or hand search*).ti,ab,kf.

20 (mantel haenszel or peto or der simonian or dersimonian or fixed effect* or latin square*).ti,ab,kf.

21 (met analy* or metanaly* or technology assessment* or HTA or HTAs or technology overview* or technology appraisal*).ti,ab,kf.

22 (meta regression* or metaregression*).ti,ab,kf.

23 (meta-analy* or metaanaly* or systematic review* or biomedical technology assessment* or bio-medical technology assessment*).mp,hw.

24 (medline or cochrane or pubmed or medlars or embase or cinahl).ti,ab,hw.

25 (cochrane or (health adj2 technology assessment) or evidence report).jw.

26 (comparative adj3 (efficacy or effectiveness)).ti,ab,kf.

27 (outcomes research or relative effectiveness).ti,ab,kf.

28 ((indirect or indirect treatment or mixed-treatment or bayesian) adj3 comparison*).ti,ab,kf.

29 (multi* adj3 treatment adj3 comparison*).ti,ab,kf.

30 (mixed adj3 treatment adj3 (meta-analy* or metaanaly*)).ti,ab,kf.

31 umbrella review*.ti,ab,kf.

32 (multi* adj2 paramet* adj2 evidence adj2 synthesis).ti,ab,kf.

33 (multiparamet* adj2 evidence adj2 synthesis).ti,ab,kf.

34 (multi-paramet* adj2 evidence adj2 synthesis).ti,ab,kf.

35 or/10-34

36 predict:.tw. or validat:.mp. or develop.tw.

37 (stratification or ROC curve).ti,ab. or exp ROC curve/ or discriminat$.ti,ab. or c-statistic.ti,ab. or "Area under the curve".ti,ab. or AUC.ti,ab. or Calibration.ti,ab. or indices.ti,ab. or algorithm.ti,ab. or multivaria$.mp.

38 Validat*.mp. or Predict$.ti. or Rule*.mp. or (Predict* and (Outcome* or Risk* or Model$)).mp. or ((History or Variable$ or Criteria or Scor$ or Characteristic$ or Finding$ or Factor$) and (Predict$ or Model$ or Decision$ or Identif$ or Prognos$)).mp. or (Decision$.mp. and ((Model$ or Clinical$).mp. or Logistic Models/)) or (Prognostic and (History or Variable$ or Criteria or Scor$ or Characteristic$ or Finding$ or Factor$ or Model$)).mp. [mp=title, book title, abstract, original title, name of substance word, subject heading word, floating sub-heading word, keyword heading word, organism supplementary concept word, protocol supplementary concept word, rare disease supplementary concept word, unique identifier, synonyms]

39 36 or 37

40 39 or 38

41 9 and 35 and 40

42 limit 41 to dt=20230101-20241231

43 limit 41 to ez=20230101-20241231

44 limit 42 to da=20230101-20241231

45   42 or 43 or 44

**Embase (OVID)**

**Date run: 31/01/23**

Database: Embase <1974 to 2023 January 30>

Search Strategy:

--------------------------------------------------------------------------------

1 (decubit* or bedsore* or bed-sore* or pressure-ulcer* or pressure-wound*).tw. (24186)

2 ((pressure* or bed or bedbound or bed-bound or bedridden or bed-ridden or deep tissue* or deep-tissue) adj3 (wound* or ulcer* or sore* or injur* or lesion*)).tw. (28692)

3 exp decubitus/ (24330)

4 1 or 2 or 3 (45109)

5 ((supine or immobil*) adj3 (heal or healing or heals or healed or dress*)).tw. (279)

6 ((supine or immobil*) adj3 (wound* or ulcer* or sore* or injur* or lesion*)).tw. (1181)

7 ((pressure or bedbound or bedridden or bed-bound or bed-ridden or deep tissue or deep-tissue) adj3 (heal or healing or heals or healed or dress*)).tw. (2463)

8 5 or 6 or 7 (3873)

9 4 or 8 (47397)

10 (systematic review or meta-analysis).pt. (0)

11 review.pt. (3006963)

12 search:.tw. (777617)

13 meta-analys:.mp. (420287)

14 meta-analysis/ or systematic review/ or systematic reviews as topic/ or meta-analysis as topic/ or "meta analysis (topic)"/ or "systematic review (topic)"/ or exp technology assessment, biomedical/ or network meta-analysis/ (600348)

15 ((systematic* adj3 (review* or overview*)) or (methodologic* adj3 (review* or overview*))).ti,ab,kf. (374423)

16 ((quantitative adj3 (review* or overview* or synthes*)) or (research adj3 (integrati* or overview*))).ti,ab,kf. (17492)

17 ((integrative adj3 (review* or overview*)) or (collaborative adj3 (review* or overview*)) or (pool* adj3 analy*)).ti,ab,kf. (53239)

18 (data synthes* or data extraction* or data abstraction*).ti,ab,kf. (47755)

19 (handsearch* or hand search*).ti,ab,kf. (13352)

20 (mantel haenszel or peto or der simonian or dersimonian or fixed effect* or latin square*).ti,ab,kf. (45672)

21 (met analy* or metanaly* or technology assessment* or HTA or HTAs or technology overview* or technology appraisal*).ti,ab,kf. (19758)

22 (meta regression* or metaregression*).ti,ab,kf. (17239)

23 (meta-analy* or metaanaly* or systematic review* or biomedical technology assessment* or bio-medical technology assessment*).mp,hw. (709507)

24 (medline or cochrane or pubmed or medlars or embase or cinahl).ti,ab,hw. (427516)

25 (cochrane or (health adj2 technology assessment) or evidence report).jw. (30592)

26 (comparative adj3 (efficacy or effectiveness)).ti,ab,kf. (25197)

27 (outcomes research or relative effectiveness).ti,ab,kf. (15991)

28 ((indirect or indirect treatment or mixed-treatment or bayesian) adj3 comparison*).ti,ab,kf. (7335)

29 (multi* adj3 treatment adj3 comparison*).ti,ab,kf. (423)

30 (mixed adj3 treatment adj3 (meta-analy* or metaanaly*)).ti,ab,kf. (256)

31 umbrella review*.ti,ab,kf. (1367)

32 (multi* adj2 paramet* adj2 evidence adj2 synthesis).ti,ab,kf. (28)

33 (multiparamet* adj2 evidence adj2 synthesis).ti,ab,kf. (21)

34 (multi-paramet* adj2 evidence adj2 synthesis).ti,ab,kf. (23)

35 or/10-34 (3937855)

36 validat:.mp. or index.tw. or model.tw. (5261274)

37 (stratification or ROC curve).ti,ab. or exp receiver operating characteristic/ or discriminat$.ti,ab. or c-statistic.ti,ab. or "Area under the curve".ti,ab. or AUC.ti,ab. or Calibration.ti,ab. or indices.ti,ab. or algorithm.ti,ab. or multivaria$.mp. (2151295)

38 Validat*.mp. or Predict$.ti. or Rule*.mp. or (Predict* and (Outcome* or Risk* or Model$)).mp. or ((History or Variable$ or Criteria or Scor$ or Characteristic$ or Finding$ or Factor$) and (Predict$ or Model$ or Decision$ or Identif$ or Prognos$)).mp. or (Decision$.mp. and ((Model$ or Clinical$).mp. or Statistical model/)) or (Prognostic and (History or Variable$ or Criteria or Scor$ or Characteristic$ or Finding$ or Factor$ or Model$)).mp. [mp=title, abstract, heading word, drug trade name, original title, device manufacturer, drug manufacturer, device trade name, keyword heading word, floating subheading word, candidate term word] (8141367)

39 36 or 37 (9046587)

40 39 or 38 (10998837)

47 9 and 35 and 40 (2060)

**Embase (OVID)**

**Date run: 20/06/24**

Database: Embase <1974 to 2024 June 20>

Search Strategy:

--------------------------------------------------------------------------------

1 (decubit* or bedsore* or bed-sore* or pressure-ulcer* or pressure-wound*).tw.

2 ((pressure* or bed or bedbound or bed-bound or bedridden or bed-ridden or deep tissue* or deep-tissue) adj3 (wound* or ulcer* or sore* or injur* or lesion*)).tw.

3 exp decubitus/

4 1 or 2 or 3

5 ((supine or immobil*) adj3 (heal or healing or heals or healed or dress*)).tw.

6 ((supine or immobil*) adj3 (wound* or ulcer* or sore* or injur* or lesion*)).tw.

7 ((pressure or bedbound or bedridden or bed-bound or bed-ridden or deep tissue or deep-tissue) adj3 (heal or healing or heals or healed or dress*)).tw.

8 5 or 6 or 7

9 4 or 8

10 (systematic review or meta-analysis).pt.

11 review.pt.

12 search:.tw.

13 meta-analys:.mp.

14 meta-analysis/ or systematic review/ or systematic reviews as topic/ or meta-analysis as topic/ or "meta analysis (topic)"/ or "systematic review (topic)"/ or exp technology assessment, biomedical/ or network meta-analysis/

15 ((systematic* adj3 (review* or overview*)) or (methodologic* adj3 (review* or overview*))).ti,ab,kf.

16 ((quantitative adj3 (review* or overview* or synthes*)) or (research adj3 (integrati* or overview*))).ti,ab,kf.

17 ((integrative adj3 (review* or overview*)) or (collaborative adj3 (review* or overview*)) or (pool* adj3 analy*)).ti,ab,kf.

18 (data synthes* or data extraction* or data abstraction*).ti,ab,kf.

19 (handsearch* or hand search*).ti,ab,kf.

20 (mantel haenszel or peto or der simonian or dersimonian or fixed effect* or latin square*).ti,ab,kf.

21 (met analy* or metanaly* or technology assessment* or HTA or HTAs or technology overview* or technology appraisal*).ti,ab,kf.

22 (meta regression* or metaregression*).ti,ab,kf.

23 (meta-analy* or metaanaly* or systematic review* or biomedical technology assessment* or bio-medical technology assessment*).mp,hw.

24 (medline or cochrane or pubmed or medlars or embase or cinahl).ti,ab,hw.

25 (cochrane or (health adj2 technology assessment) or evidence report).jw.

26 (comparative adj3 (efficacy or effectiveness)).ti,ab,kf.

27 (outcomes research or relative effectiveness).ti,ab,kf.

28 ((indirect or indirect treatment or mixed-treatment or bayesian) adj3 comparison*).ti,ab,kf.

29 (multi* adj3 treatment adj3 comparison*).ti,ab,kf.

30 (mixed adj3 treatment adj3 (meta-analy* or metaanaly*)).ti,ab,kf.

31 umbrella review*.ti,ab,kf.

32 (multi* adj2 paramet* adj2 evidence adj2 synthesis).ti,ab,kf.

33 (multiparamet* adj2 evidence adj2 synthesis).ti,ab,kf.

34 (multi-paramet* adj2 evidence adj2 synthesis).ti,ab,kf.

35 or/10-34

36 validat:.mp. or index.tw. or model.tw.

37 (stratification or ROC curve).ti,ab. or exp receiver operating characteristic/ or discriminat$.ti,ab. or c-statistic.ti,ab. or "Area under the curve".ti,ab. or AUC.ti,ab. or Calibration.ti,ab. or indices.ti,ab. or algorithm.ti,ab. or multivaria$.mp.

38 Validat*.mp. or Predict$.ti. or Rule*.mp. or (Predict* and (Outcome* or Risk* or Model$)).mp. or ((History or Variable$ or Criteria or Scor$ or Characteristic$ or Finding$ or Factor$) and (Predict$ or Model$ or Decision$ or Identif$ or Prognos$)).mp. or (Decision$.mp. and ((Model$ or Clinical$).mp. or Statistical model/)) or (Prognostic and (History or Variable$ or Criteria or Scor$ or Characteristic$ or Finding$ or Factor$ or Model$)).mp. [mp=title, abstract, heading word, drug trade name, original title, device manufacturer, drug manufacturer, device trade name, keyword heading word, floating subheading word, candidate term word] (8141367)

39 36 or 37

40 39 or 38

47 9 and 35 and 40

48 limit 47 to dc=20230101-20243112

**CINAHL PLUS (EBSCOhost)**
*SEARCH DATE 02/02/23*

S1 ( ((TI decubit* OR AB decubit*) OR (TI bedsore* OR AB bedsore*) OR (TI bed-sore* OR AB bed-sore*) OR (TI pressure-ulcer* OR AB pressure-ulcer*) OR (TI pressure-wound* OR AB pressure-wound*)) ) OR ( (((TI pressure* OR AB pressure*) OR (TI bed OR AB bed) OR (TI bedbound OR AB bedbound) OR (TI bed-bound OR AB bed-bound) OR (TI bedridden OR AB bedridden) OR (TI bed-ridden OR AB bed-ridden) OR (TI "deep tissue*" OR AB "deep tissue*") OR (TI deep-tissue OR AB deep-tissue)) N3 ((TI wound* OR AB wound*) OR (TI ulcer* OR AB ulcer*) OR (TI sore* OR AB sore*) OR (TI injur* OR AB injur*) OR (TI lesion* OR AB lesion*))) ) OR ( (MH "pressure ulcer"+) OR (MH pressure) ) Expanders - Apply equivalent subjects (28 401)

S2 ( ((TI heal OR AB heal) OR (TI healing OR AB healing) OR (TI heals OR AB heals) OR (TI healed OR AB healed) OR (TI dress* OR AB dress*))) ) OR ( (((TI supine OR AB supine) OR (TI immobil* OR AB immobil*)) N3 ((TI wound* OR AB wound*) OR (TI ulcer* OR AB ulcer*) OR (TI sore* OR AB sore*) OR (TI injur* OR AB injur*) OR (TI lesion* OR AB lesion*))) ) OR ( (((TI pressure OR AB pressure) OR (TI bedbound OR AB bedbound) OR (TI bedridden OR AB bedridden) OR (TI bed-bound OR AB bed-bound) OR (TI bed-ridden OR AB bed-ridden) OR (TI "deep tissue" OR AB "deep tissue") OR (TI deep-tissue OR AB deep-tissue)) N3 ((TI heal OR AB heal) OR (TI healing OR AB healing) OR (TI heals OR AB heals) OR (TI healed OR AB healed) OR (TI dress* OR AB dress*))) ) Expanders – Apply equivalent subjects (72 286)

S3 S1 OR S2 Expanders - Apply equivalent subjects (96245)

S4 ( (PT "systematic review" OR PT meta-analysis) ) OR ( (MH meta-analysis) OR (MH "systematic review") OR (MH "systematic reviews as topic") OR (MH "meta-analysis as topic") OR (MH "meta analysis (topic)") OR (MH "systematic review (topic)") OR (MH "technology assessment, biomedical"+) OR (MH "network meta-analysis") ) OR ( (((TI systematic* OR AB systematic* OR SU systematic*) N3 ((TI review* OR AB review* OR SU review*) OR (TI overview* OR AB overview* OR SU overview*))) OR ((TI methodologic* OR AB methodologic* OR SU methodologic*) N3 ((TI review* OR AB review* OR SU review*) OR (TI overview* OR AB overview* OR SU overview*)))) ) OR ( (((TI quantitative OR AB quantitative OR SU quantitative) N3 ((TI review* OR AB review* OR SU review*) OR (TI overview* OR AB overview* OR SU overview*) OR (TI synthes* OR AB synthes* OR SU synthes*))) OR ((TI research OR AB research OR SU research) N3 ((TI integrati* OR AB integrati* OR SU integrati*) OR (TI overview* OR AB overview* OR SU overview*)))) ) OR ( (((TI integrative OR AB integrative OR SU integrative) N3 ((TI review* OR AB review* OR SU review*) OR (TI overview* OR AB overview* OR SU overview*))) OR ((TI collaborative OR AB collaborative OR SU collaborative) N3 ((TI review* OR AB review* OR SU review*) OR (TI overview* OR AB overview* OR SU overview*))) OR ((TI pool* OR AB pool* OR SU pool*) N3 (TI analy* OR AB analy* OR SU analy*))) ) OR ( ((TI "data synthes*" OR AB "data synthes*" OR SU "data synthes*") OR (TI "data extraction*" OR AB "data extraction*" OR SU "data extraction*") OR (TI "data abstraction*" OR AB "data abstraction*" OR SU "data abstraction*")) ) OR ( ((TI handsearch* OR AB handsearch* OR SU handsearch*) OR (TI "hand search*" OR AB "hand search*" OR SU "hand search*")) ) OR ( ((TI "mantel haenszel" OR AB "mantel haenszel" OR SU "mantel haenszel") OR (TI peto OR AB peto OR SU peto) OR (TI "der simonian" OR AB "der simonian" OR SU "der simonian") OR (TI dersimonian OR AB dersimonian OR SU dersimonian) OR (TI "fixed effect*" OR AB "fixed effect*" OR SU "fixed effect*") OR (TI "latin square*" OR AB "latin square*" OR SU "latin square*")) ) OR ( ((TI "met analy*" OR AB "met analy*" OR SU "met analy*") OR (TI metanaly* OR AB metanaly* OR SU metanaly*) OR (TI "technology assessment*" OR AB "technology assessment*" OR SU "technology assessment*") OR (TI HTA OR AB HTA OR SU HTA) OR (TI HTAs OR AB HTAs OR SU HTAs) OR (TI "technology overview*" OR AB "technology overview*" OR SU "technology overview*") OR (TI "technology appraisal*" OR AB "technology appraisal*" OR SU "technology appraisal*")) ) OR ( ((TI "meta regression*" OR AB "meta regression*" OR SU "meta regression*") OR (TI metaregression* OR AB metaregression* OR SU metaregression*)) ) OR ( (meta-analy* OR metaanaly* OR "systematic review*" OR "biomedical technology assessment*" OR "bio-medical technology assessment*") ,hw. ) OR ( ((TI medline OR AB medline) OR (TI cochrane OR AB cochrane) OR (TI pubmed OR AB pubmed) OR (TI medlars OR AB medlars) OR (TI embase OR AB embase) OR (TI cinahl OR AB cinahl)) ,hw. ) Expanders - Apply equivalent subjects (237782)

S5 ( (cochrane OR ( health N2 "technology assessment") OR "evidence report") .jw. ) OR ( ((TI comparative OR AB comparative OR SU comparative) N3 ((TI efficacy OR AB efficacy OR SU efficacy) OR (TI effectiveness OR AB effectiveness OR SU effectiveness))) ) OR ( ((TI "outcomes research" OR AB "outcomes research" OR SU "outcomes research") OR (TI "relative effectiveness" OR AB "relative effectiveness" OR SU "relative effectiveness")) ) OR ( (((TI indirect OR AB indirect OR SU indirect) OR (TI "indirect treatment" OR AB "indirect treatment" OR SU "indirect treatment") OR (TI mixed-treatment OR AB mixed-treatment OR SU mixed-treatment) OR (TI bayesian OR AB bayesian OR SU bayesian)) N3 (TI comparison* OR AB comparison* OR SU comparison*)) ) OR ( ((TI multi* OR AB multi* OR SU multi*) N3 (TI treatment OR AB treatment OR SU treatment) N3 (TI comparison* OR AB comparison* OR SU comparison*)) ) OR ( (TI "umbrella review*" OR AB "umbrella review*" OR SU "umbrella review*") ) OR ( ((TI multi* OR AB multi* OR SU multi*) N2 (TI paramet* OR AB paramet* OR SU paramet*) N2 (TI evidence OR AB evidence OR SU evidence) N2 (TI synthesis OR AB synthesis OR SU synthesis)) ) OR ( ((TI multiparamet* OR AB multiparamet* OR SU multiparamet*) N2 (TI evidence OR AB evidence OR SU evidence) N2 (TI synthesis OR AB synthesis OR SU synthesis)) ) OR ( ((TI multi-paramet* OR AB multi-paramet* OR SU multi-paramet*) N2 (TI evidence OR AB evidence OR SU evidence) N2 (TI synthesis OR AB synthesis OR SU synthesis)) Expanders - Apply equivalent subjects (19108)

S6 ((TI search: OR AB search:)) Expanders - Apply equivalent subjects (123419)

S7 PT review Expanders - Apply equivalent subjects (356003)

S8 S4 OR S5 OR S6 OR S7 Expanders - Apply equivalent subjects (645360)

S9 S3 AND S8 Expanders - Apply equivalent subjects (10379)

S10 (TX validat*) or (TI index or model) or (AB index or model) Expanders - Apply equivalent subjects (1286240)

S11 TI ( stratification or "ROC curve" or discriminat" or c-statistic" or "Area under the curve" or AUC or Calibration* or indices* or algorithm* or multivaria* ) OR AB ( stratification or "ROC curve" or discriminat" or c-statistic" or "Area under the curve" or AUC or Calibration* or indices* or algorithm* or multivaria* ) Expanders - Apply equivalent subjects (294871)

S12 (MH "ROC Curve") Expanders - Apply equivalent subjects (33393)

S13 TI Validat* or Predict* or Rule* or (Predict* and (Outcome* or Risk* or Model*)) or ((History or Variable* or Criteria or Scor* or Characteristic* or Finding* or Factor*) and (Predict* or Model* or Decision* or Identif* or Prognos*)) or (Decision* and ((Model* or Clinical*) or (Prognostic and (History or Variable* or Criteria or Scor* or Characteristic* or Finding* or Factor* or Model*)) Expanders - Apply equivalent subjects (144228)

S14 AB Validat* or Predict* or Rule* or (Predict* and (Outcome* or Risk* or Model*)) or ((History or Variable* or Criteria or Scor* or Characteristic* or Finding* or Factor*) and (Predict* or Model* or Decision* or Identif* or Prognos*)) or (Decision* and ((Model* or Clinical*) or (Prognostic and (History or Variable* or Criteria or Scor* or Characteristic* or Finding* or Factor* or Model*)) Expanders - Apply equivalent subjects (1482692)

S15 (MH "Models, Statistical+") Expanders - Apply equivalent subjects (40243)

S16 S10 OR S11 OR S12 OR S13 OR S14 OR S15 Expanders - Apply equivalent subjects (2119713)

S17 S9 AND S16 (3720)

**CINAHL PLUS (EBSCOhost)**
*SEARCH DATE 21/06/24*

S1 ( ((TI decubit* OR AB decubit*) OR (TI bedsore* OR AB bedsore*) OR (TI bed-sore* OR AB bed-sore*) OR (TI pressure-ulcer* OR AB pressure-ulcer*) OR (TI pressure-wound* OR AB pressure-wound*)) ) OR ( (((TI pressure* OR AB pressure*) OR (TI bed OR AB bed) OR (TI bedbound OR AB bedbound) OR (TI bed-bound OR AB bed-bound) OR (TI bedridden OR AB bedridden) OR (TI bed-ridden OR AB bed-ridden) OR (TI "deep tissue*" OR AB "deep tissue*") OR (TI deep-tissue OR AB deep-tissue)) N3 ((TI wound* OR AB wound*) OR (TI ulcer* OR AB ulcer*) OR (TI sore* OR AB sore*) OR (TI injur* OR AB injur*) OR (TI lesion* OR AB lesion*))) ) OR ( (MH "pressure ulcer"+) OR (MH pressure) ) Expanders - Apply equivalent subjects

S2 ( ((TI heal OR AB heal) OR (TI healing OR AB healing) OR (TI heals OR AB heals) OR (TI healed OR AB healed) OR (TI dress* OR AB dress*))) ) OR ( (((TI supine OR AB supine) OR (TI immobil* OR AB immobil*)) N3 ((TI wound* OR AB wound*) OR (TI ulcer* OR AB ulcer*) OR (TI sore* OR AB sore*) OR (TI injur* OR AB injur*) OR (TI lesion* OR AB lesion*))) ) OR ( (((TI pressure OR AB pressure) OR (TI bedbound OR AB bedbound) OR (TI bedridden OR AB bedridden) OR (TI bed-bound OR AB bed-bound) OR (TI bed-ridden OR AB bed-ridden) OR (TI "deep tissue" OR AB "deep tissue") OR (TI deep-tissue OR AB deep-tissue)) N3 ((TI heal OR AB heal) OR (TI healing OR AB healing) OR (TI heals OR AB heals) OR (TI healed OR AB healed) OR (TI dress* OR AB dress*))) ) Expanders – Apply equivalent subjects

S3 S1 OR S2 Expanders - Apply equivalent subjects

S4 ( (PT "systematic review" OR PT meta-analysis) ) OR ( (MH meta-analysis) OR (MH "systematic review") OR (MH "systematic reviews as topic") OR (MH "meta-analysis as topic") OR (MH "meta analysis (topic)") OR (MH "systematic review (topic)") OR (MH "technology assessment, biomedical"+) OR (MH "network meta-analysis") ) OR ( (((TI systematic* OR AB systematic* OR SU systematic*) N3 ((TI review* OR AB review* OR SU review*) OR (TI overview* OR AB overview* OR SU overview*))) OR ((TI methodologic* OR AB methodologic* OR SU methodologic*) N3 ((TI review* OR AB review* OR SU review*) OR (TI overview* OR AB overview* OR SU overview*)))) ) OR ( (((TI quantitative OR AB quantitative OR SU quantitative) N3 ((TI review* OR AB review* OR SU review*) OR (TI overview* OR AB overview* OR SU overview*) OR (TI synthes* OR AB synthes* OR SU synthes*))) OR ((TI research OR AB research OR SU research) N3 ((TI integrati* OR AB integrati* OR SU integrati*) OR (TI overview* OR AB overview* OR SU overview*)))) ) OR ( (((TI integrative OR AB integrative OR SU integrative) N3 ((TI review* OR AB review* OR SU review*) OR (TI overview* OR AB overview* OR SU overview*))) OR ((TI collaborative OR AB collaborative OR SU collaborative) N3

((TI review* OR AB review* OR SU review*) OR (TI overview* OR AB overview* OR SU overview*))) OR ((TI pool* OR AB pool* OR SU pool*) N3 (TI analy* OR AB analy* OR SU analy*))) ) OR ( ((TI "data synthes*" OR AB "data synthes*" OR SU "data synthes*") OR (TI "data extraction*" OR AB "data extraction*" OR SU "data extraction*") OR (TI "data abstraction*" OR AB "data abstraction*" OR SU "data abstraction*")) ) OR ( ((TI handsearch* OR AB handsearch* OR SU handsearch*) OR (TI "hand search*" OR AB "hand search*" OR SU "hand search*")) ) OR ( ((TI "mantel haenszel" OR AB "mantel haenszel" OR SU "mantel haenszel") OR (TI peto OR AB peto OR SU peto) OR (TI "der simonian" OR AB "der simonian" OR SU "der simonian") OR (TI dersimonian OR AB dersimonian OR SU dersimonian) OR (TI "fixed effect*" OR AB "fixed effect*" OR SU "fixed effect*") OR (TI "latin square*" OR AB "latin square*" OR SU "latin square*")) ) OR ( ((TI "met analy*" OR AB "met analy*" OR SU "met analy*") OR (TI metanaly* OR AB metanaly* OR SU metanaly*) OR (TI "technology assessment*" OR AB "technology assessment*" OR SU "technology assessment*") OR (TI HTA OR AB HTA OR SU HTA) OR (TI HTAs OR AB HTAs OR SU HTAs) OR (TI "technology overview*" OR AB "technology overview*" OR SU "technology overview*") OR (TI "technology appraisal*" OR AB "technology appraisal*" OR SU "technology appraisal*")) ) OR ( ((TI "meta regression*" OR AB "meta regression*" OR SU "meta regression*") OR (TI metaregression* OR AB metaregression* OR SU metaregression*)) ) OR ( (meta-analy* OR metaanaly* OR "systematic review*" OR "biomedical technology assessment*" OR "bio-medical technology assessment*") ,hw. ) OR ( ((TI medline OR AB medline) OR (TI cochrane OR AB cochrane) OR (TI pubmed OR AB pubmed) OR (TI medlars OR AB medlars) OR (TI embase OR AB embase) OR (TI cinahl OR AB cinahl)) ,hw. )

S5 ( (cochrane OR ( health N2 "technology assessment") OR "evidence report") .jw. ) OR ( ((TI comparative OR AB comparative OR SU comparative) N3 ((TI efficacy OR AB efficacy OR SU efficacy) OR (TI effectiveness OR AB effectiveness OR SU effectiveness))) ) OR ( ((TI "outcomes research" OR AB "outcomes research" OR SU "outcomes research") OR (TI "relative effectiveness" OR AB "relative effectiveness" OR SU "relative effectiveness")) ) OR ( (((TI indirect OR AB indirect OR SU indirect) OR (TI "indirect treatment" OR AB "indirect treatment" OR SU "indirect treatment") OR (TI mixed-treatment OR AB mixed-treatment OR SU mixed-treatment) OR (TI bayesian OR AB bayesian OR SU bayesian)) N3 (TI comparison* OR AB comparison* OR SU comparison*)) ) OR ( ((TI multi* OR AB multi* OR SU multi*) N3 (TI treatment OR AB treatment OR SU treatment) N3 (TI comparison* OR AB comparison* OR SU comparison*)) ) OR ( (TI "umbrella review*" OR AB "umbrella review*" OR SU "umbrella review*") ) OR ( ((TI multi* OR AB multi* OR SU multi*) N2 (TI paramet* OR AB paramet* OR SU paramet*) N2 (TI evidence OR AB evidence OR SU evidence) N2 (TI synthesis OR AB synthesis OR SU synthesis)) ) OR ( ((TI multiparamet* OR AB multiparamet* OR SU multiparamet*) N2 (TI evidence OR AB evidence OR SU evidence) N2 (TI synthesis OR AB synthesis OR SU synthesis)) ) OR ( ((TI multi-paramet* OR AB multi-paramet* OR SU multi-paramet*) N2 (TI evidence OR AB evidence OR SU evidence) N2 (TI synthesis OR AB synthesis OR SU synthesis))

S6 ((TI search: OR AB search:))

S7 PT review

S8 S4 OR S5 OR S6 OR S7

S9 S3 AND S8

S10 (TX validat*) or (TI index or model) or (AB index or model)

S11 TI ( stratification or "ROC curve" or discriminat" or c-statistic" or "Area under the curve" or AUC or Calibration* or indices* or algorithm* or multivaria* ) OR AB ( stratification or "ROC curve" or discriminat" or c-statistic" or "Area under the curve" or AUC or Calibration* or indices* or algorithm* or multivaria* )

S12 (MH "ROC Curve")

S13 TI Validat* or Predict* or Rule* or (Predict* and (Outcome* or Risk* or Model*)) or ((History or Variable* or Criteria or Scor* or Characteristic* or Finding* or Factor*) and (Predict* or Model* or Decision* or Identif* or Prognos*)) or (Decision* and ((Model* or Clinical*) or (Prognostic and (History or Variable* or Criteria or Scor* or Characteristic* or Finding* or Factor* or Model*))

S14 AB Validat* or Predict* or Rule* or (Predict* and (Outcome* or Risk* or Model*)) or ((History or Variable* or Criteria or Scor* or Characteristic* or Finding* or Factor*) and (Predict* or Model* or Decision* or Identif* or Prognos*)) or (Decision* and ((Model* or Clinical*) or (Prognostic and (History or Variable* or Criteria or Scor* or Characteristic* or Finding* or Factor* or Model*))

S15 (MH "Models, Statistical+")

S16 S10 OR S11 OR S12 OR S13 OR S14 OR S15

S17 S9 AND S16

S18 (EM 20230101-20241212) OR (ZD "in process" AND RD 20230101-20241212)

S19 S17 AND S18

**EPISTEMONIKOS**

**Date run: 31/01/23**
**Date update run: 21/06/24**
For update, all searches limited by date added to database: From: 01/01/2023 To: 21/06/2024

**Search 1**

(title:(decubit* OR bedsore* OR bed-sore* OR pressure-ulcer* OR pressure-wound*) OR abstract:(decubit* OR bedsore* OR bed-sore* OR pressure-ulcer* OR pressure-wound*)) AND (title:(stratification OR "ROC curve" OR "ROC curves" OR "receiver operating characteristic" OR discriminat* OR c-statistic OR "Area under the curve" OR AUC OR Calibration OR indices OR algorithm OR multivaria* OR Validat* OR Predict* OR Rule* OR Risk* OR Model* OR Criteria OR Scor* OR Characteristic* OR Finding* OR Factor* OR Decision* OR Prognos* OR Index OR model OR prevent*) OR abstract:(stratification OR "ROC curve" OR "ROC curves" OR "receiver operating characteristic" OR discriminat* OR c-statistic OR "Area under the curve" OR AUC OR Calibration OR indices OR algorithm OR multivaria* OR Validat* OR Predict* OR Rule* OR Risk* OR Model* OR Criteria OR Scor* OR Characteristic* OR Finding* OR Factor* OR Decision* OR Prognos* OR Index OR model OR prevent*))
Limit by publication type: systematic review (106) (Update: 21) or broad synthesis (3) (Update: 0)

**Search 2**

(title:("pressure ulcer" OR "pressure ulcers" OR "pressure sore" OR "pressure sores" OR "pressure lesion" OR "pressure lesions" OR "pressure injury" OR "pressure injuries") OR abstract:("pressure

ulcer" OR "pressure ulcers" OR "pressure sore" OR "pressure sores" OR "pressure lesion" OR "pressure lesions" OR "pressure injury" OR "pressure injuries")) AND (title:(stratification OR "ROC curve" OR "ROC curves"  OR "receiver operating characteristic" OR discriminat* OR c-statistic OR "Area under the curve" OR AUC OR Calibration OR indices OR algorithm OR multivaria* OR Validat* OR Predict* OR Rule* OR Risk* OR Model* OR Criteria OR Scor* OR Characteristic* OR Finding* OR Factor* OR Decision* OR Prognos* OR Index OR model OR prevent*) OR abstract:(stratification OR "ROC curve" OR "ROC curves"  OR "receiver operating characteristic" OR discriminat* OR c-statistic OR "Area under the curve" OR AUC OR Calibration OR indices OR algorithm OR multivaria* OR Validat* OR Predict* OR Rule* OR Risk* OR Model* OR Criteria OR Scor* OR Characteristic* OR Finding* OR Factor* OR Decision* OR Prognos* OR Index OR model OR prevent*))
Limit by publication type: systematic review (709) (Update: 129) or broad synthesis (35) (Update: 25)

**Search 3**

(title:("deep-tissue wound" OR "deep-tissue wounds" OR "deep-tissue ulcer" OR "deep-tissue ulcers" OR "deep-tissue sore" OR "deep-tissue sores" OR "deep-tissue lesion" OR "deep-tissue lesions" OR "deep-tissue injury" OR "deep-tissue injuries") OR abstract:("deep-tissue wound" OR "deep-tissue wounds" OR "deep-tissue ulcer" OR "deep-tissue ulcers"  OR "deep-tissue sore" OR "deep-tissue sores" OR "deep-tissue lesion" OR "deep-tissue lesions" OR "deep-tissue injury" OR "deep-tissue injuries")) AND (title:(stratification OR "ROC curve" OR "ROC curves"  OR "receiver operating characteristic" OR discriminat* OR c-statistic OR "Area under the curve" OR AUC OR Calibration OR indices OR algorithm OR multivaria* OR Validat* OR Predict* OR Rule* OR Risk* OR Model* OR Criteria OR Scor* OR Characteristic* OR Finding* OR Factor* OR Decision* OR Prognos* OR Index OR model OR prevent*) OR abstract:(stratification OR "ROC curve" OR "ROC curves"  OR "receiver operating characteristic" OR discriminat* OR c-statistic OR "Area under the curve" OR AUC OR Calibration OR indices OR algorithm OR multivaria* OR Validat* OR Predict* OR Rule* OR Risk* OR Model* OR Criteria OR Scor* OR Characteristic* OR Finding* OR Factor* OR Decision* OR Prognos* OR Index OR model OR prevent*))
Limit by publication type: systematic review (0) (Update: 0) or broad synthesis (0) (Update: 0)

**Search 4**

(title:("deep tissue wound" OR "deep tissue wounds" OR "deep tissue ulcer" OR "deep tissue ulcers" OR "deep tissue sore" OR "deep tissue sores" OR "deep tissue lesion" OR "deep tissue lesions" OR "deep tissue injury" OR "deep tissue injuries") OR abstract:("deep tissue wound" OR "deep tissue wounds" OR "deep tissue ulcer" OR "deep tissue ulcers"  OR "deep tissue sore" OR "deep tissue sores" OR "deep tissue lesion" OR "deep tissue lesions" OR "deep tissue injury" OR "deep tissue injuries")) AND (title:(stratification OR "ROC curve" OR "ROC curves"  OR "receiver operating characteristic" OR discriminat* OR c-statistic OR "Area under the curve" OR AUC OR Calibration OR indices OR algorithm OR multivaria* OR Validat* OR Predict* OR Rule* OR Risk* OR Model* OR Criteria OR Scor* OR Characteristic* OR Finding* OR Factor* OR Decision* OR Prognos* OR Index OR model OR prevent*) OR abstract:(stratification OR "ROC curve" OR "ROC curves"  OR "receiver operating characteristic" OR discriminat* OR c-statistic OR "Area under the curve" OR AUC OR Calibration OR indices OR algorithm OR multivaria* OR Validat* OR Predict* OR Rule* OR Risk* OR Model* OR Criteria OR Scor* OR Characteristic* OR Finding* OR Factor* OR Decision* OR Prognos* OR Index OR model OR prevent*))
Limit by publication type: systematic review (5) (Update: 1) or broad synthesis (2) (Update: 0)

**Search 5**

(title:("bed wound" OR "bed wounds" OR "bed ulcer" OR "bed ulcers" OR "bed sore" OR "bed sores" OR "bed lesion" OR "bed lesions" OR "bed injury" OR "bed injuries") OR abstract:("bed wound" OR "bed wounds" OR "bed ulcer" OR "bed ulcers" OR "bed sore" OR "bed sores" OR "bed lesion" OR "bed lesions" OR "bed injury" OR "bed injuries")) AND (title:(stratification OR "ROC curve" OR "ROC curves" OR "receiver operating characteristic" OR discriminat* OR c-statistic OR "Area under the curve" OR AUC OR Calibration OR indices OR algorithm OR multivaria* OR Validat* OR Predict* OR Rule* OR Risk* OR Model* OR Criteria OR Scor* OR Characteristic* OR Finding* OR Factor* OR Decision* OR Prognos* OR Index OR model OR prevent*) OR abstract:(stratification OR "ROC curve" OR "ROC curves" OR "receiver operating characteristic" OR discriminat* OR c-statistic OR "Area under the curve" OR AUC OR Calibration OR indices OR algorithm OR multivaria* OR Validat* OR Predict* OR Rule* OR Risk* OR Model* OR Criteria OR Scor* OR Characteristic* OR Finding* OR Factor* OR Decision* OR Prognos* OR Index OR model OR prevent*))
Limit by publication type: systematic review (14) (Update: 0) or broad synthesis (0) (Update: 1)

**Search 6**

(title:("bed bound" OR bed-bound OR bedridden OR bed-ridden OR "bed ridden") OR abstract:("bed bound" OR bed-bound OR bedridden OR bed-ridden OR "bed ridden")) AND (title:(stratification OR "ROC curve" OR "ROC curves" OR "receiver operating characteristic" OR discriminat* OR c-statistic OR "Area under the curve" OR AUC OR Calibration OR indices OR algorithm OR multivaria* OR Validat* OR Predict* OR Rule* OR Risk* OR Model* OR Criteria OR Scor* OR Characteristic* OR Finding* OR Factor* OR Decision* OR Prognos* OR Index OR model OR prevent*) OR abstract:(stratification OR "ROC curve" OR "ROC curves" OR "receiver operating characteristic" OR discriminat* OR c-statistic OR "Area under the curve" OR AUC OR Calibration OR indices OR algorithm OR multivaria* OR Validat* OR Predict* OR Rule* OR Risk* OR Model* OR Criteria OR Scor* OR Characteristic* OR Finding* OR Factor* OR Decision* OR Prognos* OR Index OR model OR prevent*))
Limit by publication type: systematic review (30) (Update: 3) or broad synthesis (1) (Update: 1)

**Search 7**

(title:(stratification OR "ROC curve" OR "ROC curves" OR "receiver operating characteristic" OR discriminat* OR c-statistic OR "Area under the curve" OR AUC OR Calibration OR indices OR algorithm OR multivaria* OR Validat* OR Predict* OR Rule* OR Risk* OR Model* OR Criteria OR Scor* OR Characteristic* OR Finding* OR Factor* OR Decision* OR Prognos* OR Index OR model OR prevent*) OR abstract:(stratification OR "ROC curve" OR "ROC curves" OR "receiver operating characteristic" OR discriminat* OR c-statistic OR "Area under the curve" OR AUC OR Calibration OR indices OR algorithm OR multivaria* OR Validat* OR Predict* OR Rule* OR Risk* OR Model* OR Criteria OR Scor* OR Characteristic* OR Finding* OR Factor* OR Decision* OR Prognos* OR Index OR model OR prevent*)) AND (title:( wound* OR ulcer* OR sore* OR injur* OR lesion*) OR abstract:( wound* OR ulcer* OR sore* OR injur* OR lesion*)) AND (title:(supine OR immobil*) OR abstract:(supine OR immobil*))
Limit by publication type: systematic review (234) (Update: 39) or broad synthesis (13) (Update: 3)

**Search 8**

(title:(stratification OR "ROC curve" OR "ROC curves" OR "receiver operating characteristic" OR discriminat* OR c-statistic OR "Area under the curve" OR AUC OR Calibration OR indices OR algorithm OR multivaria* OR Validat* OR Predict* OR Rule* OR Risk* OR Model* OR Criteria OR

Scor* OR Characteristic* OR Finding* OR Factor* OR Decision* OR Prognos* OR Index OR model OR prevent*) OR abstract:(stratification OR "ROC curve" OR "ROC curves" OR "receiver operating characteristic" OR discriminat* OR c-statistic OR "Area under the curve" OR AUC OR Calibration OR indices OR algorithm OR multivaria* OR Validat* OR Predict* OR Rule* OR Risk* OR Model* OR Criteria OR Scor* OR Characteristic* OR Finding* OR Factor* OR Decision* OR Prognos* OR Index OR model OR prevent*)) AND (title:(heal OR healing OR heals OR healed OR dress*) OR abstract:(heal OR healing OR heals OR healed OR dress*)) AND (title:(supine OR immobil*) OR abstract:(supine OR immobil*))
Limit by publication type: systematic review (39) (Update: 9) or broad synthesis (3) (Update: 0)

**GOOGLE SCHOLAR 1/02/23**
**Update run: 24/06/24**, limit to years 2023-2024

allintitle: prevent OR prevention OR risk OR predict OR prevents OR risks OR prediction OR predicts OR prognosis OR prognostic "pressure injury" -ulcer -ulcers Limit to review and years 2013-2023 (66) (Update: 31)

allintitle: prevent OR prevention OR risk OR predict OR prevents OR risks OR prediction OR predicts OR prognosis OR prognostic "pressure injuries" -ulcer -ulcers Limit to review and years 2013-2023 (33)
(Update: 14)

allintitle: prevent OR prevention OR risk OR predict OR prevents OR risks OR prediction OR predicts OR prognosis OR prognostic "pressure ulcer" Limit to review and years 2013-2023 (120)
(Update: 14)

allintitle: prevent OR prevention OR risk OR predict OR prevents OR risks OR prediction OR predicts OR prognosis OR prognostic "pressure ulcers" Limit to review and years 2013-2023 (102)
(Update: 14)

allintitle: prevent OR prevention OR risk OR predict OR prevents OR risks OR prediction OR predicts OR prognosis OR prognostic "pressure sore" Limit to review and years 2013-2023 (3)
(Update: 0)

allintitle: prevent OR prevention OR risk OR predict OR prevents OR risks OR prediction OR predicts OR prognosis OR prognostic "pressure sores" Limit to review and years 2013-2023 (2)
(Update: 0)

allintitle: prevent OR prevention OR risk OR predict OR prevents OR risks OR prediction OR predicts OR prognosis OR prognostic "pressure wound" Limit to review and years 2013-2023 (25)
(Update: 4)

allintitle: prevent OR prevention OR risk OR predict OR prevents OR risks OR prediction OR predicts OR prognosis OR prognostic "pressure wounds" Limit to review and years 2013-2023 (0)
(Update: 0)

allintitle: prevent OR prevention OR risk OR predict OR prevents OR risks OR prediction OR predicts OR prognosis OR prognostic "bedsore" Limit to review and years 2013-2023 (1)
(Update: 0)

allintitle: prevent OR prevention OR risk OR predict OR prevents OR risks OR prediction OR predicts OR prognosis OR prognostic "bedsores” Limit to review and years 2013-2023 (1)
(Update: 1)

allintitle: prevent OR prevention OR risk OR predict OR prevents OR risks OR prediction OR predicts OR prognosis OR prognostic "bed sore” Limit to review and years 2013-2023 (0)
(Update: 0)

allintitle: prevent OR prevention OR risk OR predict OR prevents OR risks OR prediction OR predicts OR prognosis OR prognostic "bed sores” Limit to review and years 2013-2023 (0)
(Update: 0)

allintitle: prevent OR prevention OR risk OR predict OR prevents OR risks OR prediction OR predicts OR prognosis OR prognostic "decubitus” Limit to review and years 2013-2023 (4)
(Update: 2)

#### Appendix 3: Data extraction form

| Data Extraction Items | |
|---|---|
| | Extractor |
| **Publication information:** | Review Title;<br>First Author;<br>Publication Year;<br>Umbrella review eligibility (DTA, CE);<br>Comments;<br>Primary studies fundings reported?[A];<br>Conflicts of Interest reported?[A] |
| **Eligibility Criteria:** | Population;<br>Setting;<br>Prediction models/tools;<br>Model outcome (and classification if specified);<br>Interventions[B];<br>Comparators[B];<br>Outcomes of interest[B];<br>Inclusion criteria incorporated PICO, PIRT or POII?[A];<br>Source of data (prospective/retrospective);<br>Phase of development of models;<br>Study design;<br>Did they explain reasons for study design inclusions?[A];<br>Exclusion criteria |
| **Review methods:** | Review protocol;<br>Protocol and justifications for deviations from?[A];<br>Databases searched;<br>Adequate search strategy?[A];<br>Search cut-off date;<br>Publication restrictions;<br>Quality assessment tool;<br>Suitable quality assessment tool?;<br>Study selection method;<br>Study selection in duplicate?[A];<br>Quality assessment method;<br>Data extraction method;<br>Data extraction in duplicate?[A];<br>Synthesis method;<br>Appropriate method of statistical synthesis, if applicable?[A] |
| **Review results:** | PRISMA diagram provided?;<br>Excluded studies list (with justifications)?[A];<br>N models per review;<br>N studies per review;<br>N participants in review;<br>How were the results presented? (e.g. outcomes reported);<br>Description of included studies provided? (summary table, tabulated per study, narrative only);<br>Description of included studies adequate?[A];<br>Study quality described? (summary table, tabulated per study, narrative only);<br>Assessment of RoB satisfactory?[A];<br>Assessment of impact of RoB on synthesised results?[A];<br>Assessment of impact of RoB on review results?[A];<br>Discussion/investigation of heterogeneity?[A];<br>Models included;<br>Brief description of included studies;<br>Brief description of study quality |

| Reviews on model accuracy | Reviews on clinical effectiveness |
|---|---|
| 2x2 tables presented for each study? | |
| Cut-off points specified for each study? | |
| List Author, year of primary studies included in review | |
| Summary estimates:<br>Sensitivity (incl. n), specificity (incl. N), likelihood ratios, DOR, AUROC, predictive values<br>Summary Sensitivity (incl. n)<br>(results from statistical synthesis) | Summary of statistical synthesis of results<br>(e.g. effect on incidence of PI, treatment outcome, or other patient-relevant outcomes) |
| Summary of narrative synthesis of results | Summary of narrative synthesis of results |

DTA – diagnostic test accuracy; CE – clinical effectiveness; PICO – population, intervention, comparator, outcome; PIRT – population, index test, reference standard, target condition; POII – population, outcome, intended use, intended timing; PRISMA – Preferred Reporting Items for Systematic Reviews and Meta-Analyses; RoB – risk of bias; O/E – observed/expected; AUC – area under the curve; DOR – diagnostic odds ratio; AUROC – area under the receiver operating characteristic curve; PI – pressure injury; AMSTAR – A MeaSurement Tool to Assess systematic Reviews

[A] AMSTAR-2 Items.

[B] applicable to clinical effectiveness reviews only.

Appendix 4: AMSTAR-2 Methodology Quality Appraisal. Adapted for application to reviews of prognostic model and accuracy studies.

| | AMSTAR-2 Adapted | |
|---|---|---|
| | Questions | Guidance |
| Item 1. | 1. Did the research questions and inclusion criteria for the review include the components one of the following: PICO, or PIRT and POII?<br>**Y/N** | For intervention reviews:<br>Population, Intervention, Comparator, Outcome (PICO)<br><br>For prognostic accuracy reviews:<br>Population, Index test, Reference standard, Target condition (PIRT)<br>Population, Outcome to be predicted, Intended use of model, Intended moment in time (POII) |
| **Item 2*** | 2. Did the report of the review contain an explicit statement that the review methods were established prior to the conduct of the review and did the report justify any significant deviations from the protocol?<br>**Y/PY/N** | For Partial Yes (PY):<br>The authors state that they had a written protocol or guide that included ALL the following:<br>• review question(s),<br>• a search strategy,<br>• inclusion/exclusion criteria,<br>• a risk of bias assessment.<br>For Yes:<br>As for partial yes, plus the protocol should be registered and should also have specified:<br>• a meta-analysis/synthesis plan, if appropriate,<br>• and a plan for investigating causes of heterogeneity,<br>• justification for any deviations from the protocol. |

| | | |
|---|---|---|
| Item 3. | 3. Did the review authors explain their selection of the study designs for inclusion in the review?<br>**Y/N** | For Yes, the review should give an explanation for including types of studies included in the review, for example:<br><br>For the DEV/VAL review: development studies, validation studies or both.<br>For the accuracy/effectiveness review: single group (prospective/retrospective), two/multi group (i.e diagnostic case-control), RCTs, NSRs |
| **Item 4*** | 4. Did the review authors use a comprehensive literature search strategy?<br>**Y/PY/N** | For Partial Yes (all the following):<br>• searched at least 2 databases (relevant to research question),<br>• provided key word and/or search strategy,<br>• justified publication restrictions (e.g. language).<br>For Yes, should also have (all the following):<br>• searched the reference lists / bibliographies of included studies,<br>• searched trial/study registries,<br>• included/consulted content experts in the field where relevant,<br>• searched for grey literature,<br>• conducted search within 24 months of completion of the review. |
| Item 5. | 5. Did the review authors perform study selection in duplicate?<br>**Y/N** | For Yes, either ONE of the following:<br>at least two reviewers independently agreed on selection of eligible studies and achieved consensus on which studies to include,<br>OR two reviewers selected a sample of eligible studies and achieved good agreement (at least 80 percent), with the remainder selected by one reviewer. |
| Item 6. | 6. Did the review authors perform data extraction in duplicate?<br>**Y/N** | For Yes, either ONE of the following:<br>at least two reviewers achieved consensus on which data to extract from included studies,<br>OR two reviewers extracted data from a sample of eligible studies and achieved good agreement (at least 80 percent), with the remainder extracted by one reviewer. |

| | | |
|---|---|---|
| **Item 7*** | 7. Did the review authors provide a list of excluded studies and justify the exclusions?<br>**Y/PY/N** | For Partial Yes:<br>provided a list of all potentially relevant studies that were read in full-text form but excluded from the review<br><br>For Yes, must also have:<br>Justified the exclusion from the review of each potentially relevant study |
| Item 8. | 8. Did the review authors describe the included studies in adequate detail?<br>**Y/PY/N** | For Partial Yes (ALL the following per included study):<br>• described PICO/PIRT/POII (whichever applicable),<br>• and described research designs<br>For Yes, should also have ALL the following per included study:<br>• described PICO/PIRT/POII (whichever applicable) in detail,<br>• described study's setting<br>• and timeframe for follow-up |
| **Item 9*** | 9. Did the review authors use a satisfactory technique for assessing the risk of bias (RoB) in individual studies that were included in the review?<br>**Y/PY/N** | **RCTs**<br>For Partial Yes, must have reported summary findings and assessed RoB from:<br>• unconcealed allocation,<br>• and lack of blinding of patients and assessors when assessing outcomes (unnecessary for objective outcomes such as all-cause mortality)<br>For Yes, must also have given itemisation of quality judgements per study, and assessed RoB from:<br>• allocation sequence that was not truly random,<br>• and selection of the reported result from among multiple measurements or analyses of a specified outcome |
| | | **NRS**<br>For Partial Yes, must have reported summary findings and assessed RoB from:<br>• confounding,<br>• and from selection bias<br>For Yes, must also have given itemisation of quality judgements per study, and assessed RoB from:<br>• methods used to ascertain exposures and outcomes,<br>• and selection of the reported result from among multiple measurements or analyses of a specified outcome |

| | | |
|---|---|---|
| | | **DTA studies**<br>For Partial Yes, must have assessed RoB with a recognised tool (e.g. QUADAS-2) and given summary of result across domains<br><br>For Yes, must also have also given itemisation of quality judgements per study. |
| | | **Prognostic studies**<br>For Partial Yes, must have assessed RoB with a recognised tool (e.g. PROBAST, QUIPS) and given summary of result across domains<br><br>For Yes, must also have also given itemisation of quality judgements per study. |
| Item 10. | 10. Did the review authors report on the sources of funding for the studies included in the review?<br>**Y/N** | For Yes:<br>Must have reported on the sources of funding for individual studies included in the review. Note: Reporting that the reviewers looked for this information, but it was not reported by study authors also qualifies |
| **Item 11*** | 11. If meta-analysis was performed did the review authors use appropriate methods for statistical combination of results?<br>**Y/N/ 'No MA conducted'** | For Yes:<br>The authors justified combining the data in a meta-analysis<br>AND they used an appropriate weighted technique to combine study results and adjusted for heterogeneity if present.<br>AND investigated the causes of any heterogeneity |
| Item 12. | 12. If meta-analysis was performed, did the review authors assess the potential impact of RoB in individual studies on the results of the meta-analysis or other evidence synthesis?<br>**Y/N/ 'No MA conducted'** | For Yes:<br>included only low risk of bias studies<br>OR, if the pooled estimate was based on studies at variable RoB, the authors performed sensitivity analyses to investigate possible impact of RoB on summary estimates |
| **Item 13*** | 13. Did the review authors account for RoB in individual studies when interpreting/ discussing the results of the review?<br>**Y/N** | For Yes:<br>included only low risk of bias RCTs<br>OR, if RCTs with moderate or high RoB, or NRSs were included the review provided a discussion of the likely impact of RoB on the results |

| | | |
|---|---|---|
| Item 14. | 14. Did the review authors provide a satisfactory explanation for, and discussion of, any heterogeneity observed in the results of the review?<br>**Y/N** | For Yes:<br>There was no significant heterogeneity,<br>OR if heterogeneity was present, the authors performed an investigation of main sources of heterogeneity in the results, if applicable, and particularly any between-study heterogeneity and discussed the impact of this |
| Item 15. | 15. Did the review authors report any potential sources of conflict of interest, including any funding they received for conducting the review?<br>**Y/N** | For Yes:<br>The authors reported no competing interests,<br>OR the authors described their funding sources and how they managed potential conflicts of interest |

* Critical domains identified by AMSTAR-2 developers.[1]

PICO – population, intervention, comparator, outcome; PIRT – population, index test, reference standard, target condition; POII – population, outcome, intended use, intended time; DEV/VAL – development/validation; RCT – randomised controlled trial; NRS – non-randomised study; QUADAS – Quality Assessment of Diagnostic Accuracy Studies; PROBAST – Prediction model Risk Of Bias ASsessment Tool; QUIPS – Quality In Prognosis Studies; MA – Meta-Analysis

#### Appendix 5: Detailed results tables

##### Table A1. Full-text articles excluded, with reasons

| | Author, year | Title | Major reason for exclusion |
|---|---|---|---|
| 1 | Alves, 2014 [2] | *Assessment of risk for pressure ulcers in intensive care units: an integrative review* | Not a systematic review |
| 2 | Anthony, 2008 [3] | *Norton, Waterlow and Braden scores: a review of the literature and a comparison between the scores and clinical judgement* | Not a systematic review |
| 3 | Barradas Cavalcante, 2016 [4] | *Updating pf the assistance protocol for pressure prevention: evidence based practice* | Not a systematic review |
| 4 | Charalambous, 2018 [5] | *Evaluation of the Validity and Reliability of the Waterlow Pressure Ulcer Risk Assessment Scale* | Not a systematic review |
| 5 | de Laat, 2006 [6] | *Epidemiology, risk and prevention of pressure ulcers in critically ill patients: a literature review* | Not a systematic review |
| 6 | do Egito Cavalcanti de Farias, 2022 [7] | *Risk factors for the development of pressure injury in the elderly: integrative review* | Not a systematic review |
| 7 | Feuchtinger, 2005 [8] | *Pressure ulcer risk factors in cardiac surgery: A review of the research literature* | Not a systematic review |
| 8 | Garcia-Fernandez, 2014 [9] | *A new theoretical model for the development of pressure ulcers and other dependence-related lesions* | Not a systematic review |
| 9 | Garrubba, 2017 [10] | *Effectiveness of the Braden risk screening tool for pressure injuries: systematic review* | Not a systematic review |
| 10 | Kelechi, 2013 [11] | *Review of pressure ulcer risk assessment scales* | Not a systematic review |
| 11 | Keller, 2002 [12] | *Pressure ulcers in intensive care patients: A review of risks and prevention* | Not a systematic review |
| 12 | Ladd, 2018 [13] | *A systematic review of pressure ulcers in burn patients: Risk factors, demographics, and treatment modalities* | Not a systematic review |
| 13 | Lepisto, 2006 [14] | *Developing a Pressure Ulcer Risk Assessment Scale for Patients in Long-Term Care* | Not a systematic review |
| 14 | Mendes Coqueiro, 2013 [15] | *Multiple risk factors and preventive strategies of pressure ulcers: systematic review* | Not a systematic review |
| 15 | Michel, 2012 [16] | *As of 2012, what are the key predictive risk factors for pressure ulcers? Developing French guidelines for clinical practice* | Not a systematic review |
| 16 | Ming, 2012 [17] | *Systematic review of pressure ulcer risk assessment scales for using in ICU patients* | Not a systematic review |
| 17 | Mordiffi, 2010 [18] | *Evaluating the effects of using the mobility assessment sub-scale within the Braden Scale on pressure ulcer incidence and preventive interventions in adult acute care settings: A systematic review* | Not a systematic review |
| 18 | Mordiffi, 2011 [19] | *Use of mobility subscale for risk assessment of pressure ulcer incidence and preventive interventions: A systematic review* | Not a systematic review |
| 19 | Mortenson, 2008 [20] | *A review of scales for assessing the risk of developing a pressure ulcer in individuals with SCI* | Not a systematic review |
| 20 | Nadeem, 2021 [21] | *Utility of the Waterlow scale in acute care settings: A literature review* | Not a systematic review |
| 21 | O'Tuathail, 2011 [22] | *Evaluation of three commonly used pressure ulcer risk assessment scales* | Not a systematic review |
| 22 | Rodriguez Tores, 2007 [23] | *Clinical judgement or assessment scales to identify patients at risk of developing pressure ulcers?* | Not a systematic review |
| 23 | Sales de Almeida, 2020 [24] | *Pressure injury prevention scales in intensive care units: an integrative review* | Not a systematic review |
| 24 | Santos, 2015 [25] | *Development of the nursing diagnosis risk for pressure ulcer* | Not a systematic review |
| 25 | Satekova, 2014 [26] | *Validity of pressure ulcer risk assesment scales: Review* | Not a systematic review |
| 26 | Shahin, 2007 [27] | *Predictive validity of pressure ulcer risk assessment tools in intensive care patients* | Not a systematic review |

| | | | |
|---|---|---|---|
| 27 | Smet, 2019 [28] | *The Belgian pressure ulcer risk assessment project: Is assessing mobility and skin status a more accurate, reliable, and feasible approach to assess pressure ulcer risk in hospitalised patients?* | Not a systematic review |
| 28 | Solati, 2016 [29] | *Predictive values of Braden and Waterlow scales to assess the risk of pressure ulcer* | Not a systematic review |
| 29 | Taylor, 1988 [30] | *Assessment tools for the identification of patients at risk for the development of pressure sores: a review* | Not a systematic review |
| 30 | Tran, 2016 [31] | *Prevention of Pressure Ulcers in the Acute Care Setting: New Innovations and Technologies* | Not a systematic review |
| 31 | Tschannen, 2020 [32] | *The pressure injury predictive model: A framework for hospital-acquired pressure injuries* | Not a systematic review |
| 32 | Walsh, 2011 [33] | *Investigating the reliability and validity of the Waterlow risk assessment scale: A literature review* | Not a systematic review |
| 33 | Xu, 2018 [34] | *Risk assessment tools for pressure injury in intensive care patients: a review* | Not a systematic review |
| 34 | Alderden, 2017 [35] | *Risk factors for pressure injuries among critical care patients: A systematic review* | No risk prediction models |
| 35 | Barbosa da Silva, 2020 [36] | *Pressure ulcers in individuals with spinal cord injury: risk factors in neurological rehabilitation* | No risk prediction models |
| 36 | Di Prinzio, 2019 [37] | *Risk factors for the development and recurrence of pressure ulcers in patients with spinal cord injury: A systematic review* | No risk prediction models |
| 37 | Haisley, 2020 [38] | *Postoperative pressure injuries in adults having surgery under general anaesthesia: systematic review of perioperative risk factors* | No risk prediction models |
| 38 | Ham, 2014 [39] | *Pressure ulcers from spinal immobilization in trauma patients: A systematic review* | No risk prediction models |
| 39 | Lima, 2021 [40] | *Risk factors and preventive interventions for pressure injuries in cancer patients* | No risk prediction models |
| 40 | Lima Serrano, 2017 [41] | *Risk factors for pressure ulcer development in Intensive Care Units: Systematic review* | No risk prediction models |
| 41 | Marin, 2013 [42] | *A systematic review of risk factors for the development and recurrence of pressure ulcers in people with spinal cord injuries* | No risk prediction models |
| 42 | Rao, 2016 [43] | *Risk Factors Associated With Pressure Ulcer Formation in Critically Ill Cardiac Surgery Patients: A Systematic Review* | No risk prediction models |
| 43 | Reenalda, 2009 [44] | *Clinical use of interface pressure to predict pressure ulcer development: a systematic review* | No risk prediction models |
| 44 | Shi, 2018 [45] | *Skin status for predicting pressure ulcer development: A systematic review and meta-analyses* | No risk prediction models |
| 45 | Siping, 2022 [46] | *Risk factors of intraoperative acquired pressure injury: A systematic review and meta-analysis* | No risk prediction models |
| 46 | Wynn, 2022 [47] | *Risk factors for the development and evolution of deep tissue injuries: A systematic review* | No risk prediction models |
| 47 | Zhang, 2022 [48] | *Prevalence and Risk Factors of Postoperative Pressure Ulcers: A Systematic Review and Meta-analysis of Diagnostic Test* | No risk prediction models |
| 48 | Bulfone, 2018 [49] | *Perioperative Pressure Injuries: A Systematic Literature Review* | Wrong research question |
| 49 | Chung, 2022 [50] | *Risk Factors for Pressure Injuries in Adult Patients: A Narrative Synthesis* | Wrong research question |
| 50 | Chung, 2022 [51] | *Risk factors for pressure ulcers in adult patients: A meta-analysis on sociodemographic factors and the Braden scale* | Wrong research question |
| 51 | Coleman, 2013 [52] | *Patient risk factors for pressure ulcer development: Systematic review* | Wrong research question |
| 52 | Dube, 2022 [53] | *Risk factors associated with heel pressure ulcer development in adult population: A systematic literature review* | Wrong research question |
| 53 | Ferris, 2019 [54] | *Pressure ulcers in patients receiving palliative care: A systematic review* | Wrong research question |
| 54 | Floyd, 2018 [55] | *Effectiveness of pressure ulcer protocols with the Braden Scale for elderly patients in the intensive care unit: A Systematic Review* | Wrong research question |
| 55 | Gelis, 2009 [56] | *Pressure ulcer risk factors in persons with SCI: Part I: Acute and rehabilitation stages* | Wrong research question |

| | | | |
|---|---|---|---|
| 56 | Gelis, 2009 [57] | *Pressure ulcer risk factors in persons with spinal cord injury part 2: the chronic stage* | Wrong research question |
| 57 | Liu, 2024 [58] | *Effects of predictive nursing interventions on pressure ulcer in older bedridden patients: A meta-analysis* | Wrong research question |
| 58 | Moore, 2023 [59] | *A systematic review of movement monitoring devices to aid the prediction of pressure ulcers in at-risk adults* | Wrong research question |
| 59 | Nixon, 2015 [60] | *Pressure UlceR Programme Of reSEarch (PURPOSE): using mixed methods (systematic reviews, prospective cohort, case study, consensus and psychometrics) to identify patient and organisational risk, develop a risk assessment tool and patient-reported outcome Quality of Life and Health Utility measures* | Wrong research question |
| 60 | Richardson, 2015 [61] | *Part 1: Pressure ulcer assessment - the development of Critical Care Pressure Ulcer Assessment Tool made Easy (CALCULATE)* | Wrong research question |
| 61 | Teixeira, 2022 [62] | *Risk factors for pressure injury in critically ill polytraumatized patients: A systematic review* | Wrong research question |
| 62 | Ting, 2021 [63] | *E-Health Decision Support Technologies in the Prevention and Management of Pressure Ulcers: A Systematic Review* | Wrong research question |
| 63 | Toffaha, 2023 [64] | *Leveraging artificial intelligence and decision support systems in hospital-acquired pressure injuries prediction: A comprehensive review* | Wrong research question |
| 64 | Fuentelsaz Gallego, 2005 [65] | *Review of literature on pressure ulcers in people aged 65 or over* | No English language translation |
| 65 | Garcia-Fernandez, 2013 [66] | *Risk assessment scales for pressure ulcer in intensive care units: A systematic review with metaanalysis* | No English language translation |
| 66 | Kottner, 2008 [67] | *Interrater reliability of the Braden scale* | No English language translation |
| 67 | Nunes de Sousa, 2023 [68] | *SCALES USED TO MEASURE PRESSURE INJURY RISK IN HOSPITALIZED PATIENTS: A REVIEW* | No English language translation |
| 68 | Pancorbo-Hidalgo, 2008 [69] | *Pressure ulcers risk assessment: clinical practice in Spain and a meta-analysis of scales effectiveness* | No English language translation |
| 69 | Park, 2014 [70] | *Predictive validity of the Braden Scale for pressure ulcer risk: a meta-analysis* | No English language translation |
| 70 | Yang, 2019 [71] | *Predictive validity of the Munro Scale for pressure injuries in surgical patients: A meta-analysis* | No English language translation |
| 71 | Barghouthi, 2023 [72] | *Systematic Review for Risks of Pressure Injury and Prediction Models Using Machine Learning Algorithms* | Reports model development or validation only |
| 72 | Dweekat, 2023 [73] | *Machine Learning Techniques, Applications, and Potential Future Opportunities in Pressure Injuries (Bedsores) Management: A Systematic Review* | Reports model development or validation only |
| 73 | Jiang, 2021 [74] | *Using Machine Learning Technologies in Pressure Injury Management: Systematic Review* | Reports model development or validation only |
| 74 | Ribeiro, 2021 [75] | *Literature review of machine-learning algorithms for pressure ulcer prevention: Challenges and opportunities* | Reports model development or validation only |
| 75 | Shi, 2019 [76] | *Evaluating the development and validation of empirically-derived prognostic models for pressure ulcer risk assessment: A systematic review* | Reports model development or validation only |
| 76 | Zhou, 2022 [77] | *A systematic review of predictive models for hospital-acquired pressure injury using machine learning* | Reports model development or validation only |
| 77 | De Queiroz, 2022 [7] | *Risk factors for the development of pressure injury in the elderly: integrative review/Fatores de risco o para desenvolvimento de lesão por pressão em idosos: revisão integrativa* | Duplicate |
| 78 | Garcia-Fernandez, 2013 [66] | *Risk assessment scales for pressure ulcers in intensive care units: A systematic review with meta-analysis* | Duplicate |
| 79 | Nixon, 2015 [60] | *Pressure UlceR Programme Of reSEarch (PURPOSE): using mixed methods (systematic reviews, prospective cohort, case study, consensus and psychometrics) to identify patient and* | Duplicate |

| | | | |
|---|---|---|---|
| | | *organisational risk, develop a risk assessment tool and patient-reported outcome Quality of Life and Health Utility measures* | |
| 80 | Nayar, 2021 [78] | *Waterlow score for risk assessment in surgical patients: a systematic review* | Wrong outcome |
| 81 | Zahia, 2020 [79] | *Pressure injury image analysis with machine learning techniques: A systematic review on previous and possible future methods* | Wrong outcome |
| 82 | Moore, 2008 [80] | *Risk assessment tools for the prevention of pressure ulcers* | Updated version included |
| 83 | Moore, 2014 [81] | *Risk assessment tools for the prevention of pressure ulcers* | Updated version included |
| 84 | Liao, 2018 [82] | *Predictive accuracy of the Braden Q Scale in risk assessment for paediatric pressure ulcer: A meta-analysis* | Wrong population |
| 85 | Ribeiro, 2013 [83] | *How effective is the development of skin care in critically ill patients using the Braden Scale scores aiming to prevent the incidence of pressure ulcers? Sistematic Literature Review* | No results |

#### Table A2. Systematic review characteristics

| Review author (publication year)<br><br>Review question | Eligibility criteria | | | Review methods | | | | Volume of evidence | |
|---|---|---|---|---|---|---|---|---|---|
| | **Population; setting** | **Prediction tools; PI classification system** | **Study design** | **Databases searched** | **Publication restrictions** (year; language; publication type) | **Quality assessment tool** | **Meta-analysis included; method of meta-analysis** | **N relevant studies in review** (n participants) | **N tools included** |
| Baris[84] (2015)<br><br>Effectiveness | Turkish populations only; NS | Braden; NS | NS | Turkish MEDLINE; PubMed; ScienceDirect; Google Scholar; YOK Thesis Search; Reference Directory of Turkey; Medicine Directory of Turkish Clinics; ULAKBIM National Database; National Library Bibliography of Turkish Articles | 1998-2012; English, Turkish; NS | None | No | 16 (2273[a]) | 2 |
| Chen[85] (2023)<br><br>Accuracy | Patients with a critical illness; ICU | Cubbin & Jackson; NS | Diagnostic studies (presenting TP, FP, TN and FN results) with any research design | EBSCO, PubMed, Ovid, Web of Science and Cochrane databases, Wangfang Data and China National Knowledge Infrastructure | Database inception - 2021; NS; reviews and expert opinions excluded | QUADAS-II | Yes; meta-analysis method unclear, SROC analysis | 9 (7684) | 1 |
| Chen[86] (2016)<br><br>Accuracy | NS; long-term care | Braden; NS | NS | PubMed; Web of Science | Inception-2015; English; NS | QUADAS | Yes; DerSimonian and Laird random-effects model, SROC analysis | 8 (41489) | 1 |

| Review author (publication year)<br><br>Review question | Eligibility criteria | | | Review methods | | | | Volume of evidence | |
|---|---|---|---|---|---|---|---|---|---|
| | **Population; setting** | **Prediction tools; PI classification system** | **Study design** | **Databases searched** | **Publication restrictions** (year; language; publication type) | **Quality assessment tool** | **Meta-analysis included; method of meta-analysis** | **N relevant studies in review** (n participants) | **N tools included** |
| Chou[87] (2013)<br><br>Accuracy Effectiveness | Adults (age ≥18y); acute care hospital, long-term and rehabilitation facilities, operative and postoperative, community (home care and wheelchair users) | PI risk assessment tools; NS | KQ1[b]: controlled or comparative randomised and nonrandomised trials, controlled or comparative observational studies<br>KQ2[b]: prospective studies of predictive validity (case-control excluded) | MEDLINE; CINAHL; Cochrane Library; grant databases; clinical trial registries | 1946-2021 (MEDLINE), 1988-2012 (CINAHL), inception- 2012 (Cochrane library); English; conference abstracts excluded | Criteria consistent with AHRQ Methods Guide for Effectiveness and Comparative Effectiveness Reviews | No; presented median accuracy results | KQ1[b]: 3<br>KQ2[b]: 47 | KQ1[b]: 4<br>KQ2[b]: 20 |
| Garcia-Fernandez[88] (2014)<br><br>Accuracy | No PIs at baseline; NS | PI risk assessment tools; NS | Controlled clinical trials, prospective cohort | Cochrane Library; Center for Reviews and Dissemination University of York; LILACS; CUIDEN Plus; Spanish Medical Index | 1962-2010; no restriction; peer-reviewed journal article | CASP for RCT/cohort studies | Yes; random-effects model | 70 (30327) | 28 |
| Gaspar[89] (2019)<br><br>Effectiveness | Adult inpatients; hospital wards or any acute unit | PI prevention strategies; NS | Prospective or retrospective; cross-sectional, comparative, pre-test and post-test, quasi-experimental, experimental, RCT, mixed-method | MEDLINE; CINAHL; PubMed; Web of Science; EBSCO Nursing & Allied Health; Cochrane Central Register of Controlled Trials; Library, Information Science & Technology Abstracts; MedicLatina | 2009-2018; English, French, Portuguese, Spanish; peer-reviewed | Evidence-Based Librarianship Critical Appraisal checklist | No | 1 (1231) | 2 |

| Review author (publication year)<br><br>Review question | Eligibility criteria | | | Review methods | | | | Volume of evidence | |
|---|---|---|---|---|---|---|---|---|---|
| | **Population; setting** | **Prediction tools; PI classification system** | **Study design** | **Databases searched** | **Publication restrictions** (year; language; publication type) | **Quality assessment tool** | **Meta-analysis included; method of meta-analysis** | **N relevant studies in review** (n participants) | **N tools included** |
| He[90] (2012)<br><br>Accuracy | NS; surgical | Braden; NS | Studies assessing predictive validity | PubMed; Web of Science | Not stated-2011; NS; NS | QUADAS | Yes; DerSimonian and Laird random-effects model, SROC analysis | 3 (609) | 1 |
| Health Quality Ontario[91] (2009)<br><br>Effectiveness | Any population at risk of developing PIs; NS | PI risk assessment tools; NS | Systematic reviews, RCTs, non-randomised controlled clinical trials | MEDLINE; MEDLINE In-Process; CINAHL; EMBASE; Cochrane Library; other non-indexed citations; | 1997-2008; English; NS | Criteria name not given | No | 3 (528) | 3 |
| Huang[92] (2021)<br><br>Accuracy | Inpatients aged ≥18y, no PIs at admission; NS | Braden; accepted standards (NPUAP, EPUAP, AHCPR, ICD-9, Bergstrom, others) | Cross-sectional, cohort | PubMed; CINAHL; EMBASE; Web of Science; Cochrane Library; bibliographies | Inception-2020; NS; NS | QUADAS-II | Yes; bivariate model, SROC analysis | 60 (49326) | 1 |
| Kottner[93] (2009)<br><br>Effectiveness (reliability) | NS; NS | Waterlow; NS | Inter- and intra-rater reliability and agreement | MEDLINE; EMBASE; CINAHL | 1985-2008; English, German; original research | Own criteria | No | 8 | 2 |
| Lovegrove[94] (2021)<br><br>Effectiveness | Adults (age ≥18y); acute hospital care | PI risk assessment tools; NS | Primary research | MEDLINE; EMBASE; EBSCO CINAHL; EBSCO; Scopus; Web of Science | 2010-2020; English; conference abstracts, posters excluded | JBI tools or analytical cross-sectional study appraisal checklist | No | 5 (1910) | 5 |
| Lovegrove[95] (2018)<br><br>Effectiveness | Adults; hospital or acute care | PI risk assessment; NS | Primary research | MEDLINE; CINAHL; Scopus; Web of Science | 2007-2017; English; non-research publications excluded | JBI tools | No | 20 | 5[b] |

| Review author (publication year)<br><br>Review question | Eligibility criteria | | | Review methods | | | | Volume of evidence | |
|---|---|---|---|---|---|---|---|---|---|
| | **Population; setting** | **Prediction tools; PI classification system** | **Study design** | **Databases searched** | **Publication restrictions** (year; language; publication type) | **Quality assessment tool** | **Meta-analysis included; method of meta-analysis** | **N relevant studies in review** (n participants) | **N tools included** |
| Mehicic[96] (2024)<br><br>Accuracy<br>Effectiveness (reliability, measurement error and convergent validity) | Adults (age ≥18y); ICU<br><br>For reliability assessment: sample of nurse-raters required | Braden; NS | Primary quantitative or mixed-methods research studies | CINAHL, EMBASE, MEDLINE, Scopus and Web of Science | Database inception - 2023; English language; Peer-reviewed | COSMIN RoB checklist | No | 34 (59325) | 1 |
| Moore[97] (2019)<br><br>Effectiveness | People without PIs, any age; any healthcare setting | PI risk assessment tools; validated PI staging system | RCTs or cluster-RCTs | MEDLINE; EMBASE; CINAHL; Cochrane Wounds Specialised Register; Cochrane Central Register of Controlled Trials | Start date between 1937-1974, until 2018; no restrictions; no restrictions | Cochrane RoB tool | No | 2 (1487) | 3 |
| Pancorbo-Hidalgo[98] (2006)<br><br>Accuracy<br>Effectiveness | No PIs at baseline; NS | PI risk assessment tools; NS | Controlled clinical trials, prospective cohort | MEDLINE; CINAHL; EBSCO; ScienceDirect; Current contents; DARE; Indice medico espanol; LILACS; CUIDEN; Cochrane Library; Springer; InterSciencia; ProQuest; Pascal | 1966-2003; Spanish, English, French, Portuguese; no restrictions | CASP Guide for clincial trials; critical assessment guide for PI assessment and prevention for cohort studies | Yes; weighted average values using inverse of variance for weights (for accuracy measures), DerSimonian and Laird random-effects model (for OR) | 33 | 13 |
| Park[99] (2016a)<br><br>Accuracy | NS; NS | Modified Braden, Waterlow, Norton, Cubbin & Jackson; NPUAP, EPUAP, AHCPR, Torrence Developmental | NS | MEDLINE; EMBASE; CINAHL; Cochrane Library; KoreaMed; NDSL; KERIS | NS-2013; NS; NS | QUADAS-II | Yes; random-effects model, SROC analysis | 17 (6143) | 5 |

| Review author (publication year)<br><br>Review question | Eligibility criteria | | | Review methods | | | | Volume of evidence | |
|---|---|---|---|---|---|---|---|---|---|
| | **Population; setting** | **Prediction tools; PI classification system** | **Study design** | **Databases searched** | **Publication restrictions** (year; language; publication type) | **Quality assessment tool** | **Meta-analysis included; method of meta-analysis** | **N relevant studies in review** (n participants) | **N tools included** |
| | | Classification of Pressure Sore | | | | | | | |
| Park[100] (2016b)<br><br>Accuracy | Elderly (age ≥60y); NS | Braden, Waterlow, Norton; NS | NS | MEDLINE; EMBASE; CINAHL; Cochrane database; KoreaMed | 1966-2013; NS; NS | QUADAS-II | Yes; random-effects model, SROC analysis | 29 (11729) | 3 |
| Park[101] (2015)<br><br>Accuracy | Adults (age ≥18y) with no PIs at baseline; hospitalised | Braden; NPUAP, AHCPR, others | Prospective | MEDLINE; EMBASE; CINAHL; KoreaMed; Cochrane Library; National Digital Science Library; Korea Education and Research Information Service | NS-2013; NS; NS | QUADAS-II | Yes; random-effects model, SROC analysis | 21 (6070) | 1 |
| Pei[102] (2023)<br><br>Accuracy | Adult; hospital | ML (if ≥1 model per study, only the *'best'* was included); NS | NS | PubMed, Embase, Cochrane Library, Web of Science, CINAHL, Grey literature and "other databases" | Database inception - 2022; English and Chinese; peer-reviewed articles or full-length conference proceedings | PROBAST | Yes; random-effects model, SROC analysis | 18 (408504) | 18 |
| Qu[103] (2022)<br><br>Accuracy | Adults with no PIs at baseline; hospital inpatients | ML; Munoz and Posthauer (2021) PI stage or as defined by the study authors | Diagnostic trials, crossover trials, cluster-controlled trials | MEDLINE; EMBASE; EBSCO; Web of Science | Start date between 1985-2010, until 2021; English; NS | QUADAS-II; PROBAST (but did not present PROBAST results) | Yes; fixed-effects or random-effects model dependent on heterogeneity assessment, ANOVA model for Bayesian network meta- | 24 (221541) | 24 |

| Review author (publication year)<br><br>Review question | Eligibility criteria | | | Review methods | | | | Volume of evidence | |
|---|---|---|---|---|---|---|---|---|---|
| | **Population; setting** | **Prediction tools; PI classification system** | **Study design** | **Databases searched** | **Publication restrictions** (year; language; publication type) | **Quality assessment tool** | **Meta-analysis included; method of meta-analysis** | **N relevant studies in review** (n participants) | **N tools included** |
| | | | | | | | analysis for diagnostic test accuracy | | |
| Tayyib[104] (2013)<br><br>Accuracy<br>Effectiveness | Adults; ICU | NS; NPUAP/EPUAP | Quantitative | MEDLINE; PubMed; CINHAL; EBSCOHost; Cochrane Library; ProQuest; Google Scholar | 2000-2012; English; journals, books, handbooks, abstracts | None | No | 11 (2119) | 9 |
| Wang[105] (2022)<br><br>Accuracy | Any age; any healthcare setting | NS; NS | Primary research and sample size, except case reports or case series | PubMed; EMBASE; CINAHL; Cochrane Library | Inception-2021; English; NS | JBI tools; NOS | Yes; fixed-effects or random-effects model dependent on heterogeneity assessment | 2 (992) | 2 |
| Wei[106] (2020)<br><br>Accuracy | Adults (age >18y); ICU | Braden; NS | NS | PubMed; Web of Science; Cochrane Library; SinoMed; CNKI; Wanfang | NS-2019; no restrictions; NS | QUADAS-II | Yes; DerSimonian and Laird random-effects model | 11 (10044) | 1 |
| Wilchesky[107] (2015)<br><br>Accuracy | NS; long-term care | Braden; NS | NS | MEDLINE; PubMed; EMBASE; PsychINFO | 1985-2013; English; journal articles (reviews and opinion papers excluded) | None | Yes; DerSimonian and Laird random-effects model | 9 (40361) | 1 |
| Zhang[108] (2021)<br><br>Accuracy | Inpatient aged >18y; ICU (stay >24h) | PI risk assessment tools; standard for judging the occurrence of PI had to be described | Cohort, case-control | PubMed/MEDLINE; EMBASE; CINAHL; Web of Science; Cochrane Library; China Biomedical Literature Service System; VIP Database; CNKI | Inception-2019; no restrictions; NS | QUADAS-II | Yes; hierarchal SROC model | 23 (15199) | 15 |

| Review author (publication year)<br><br>Review question | Eligibility criteria | | | Review methods | | | | Volume of evidence | |
|---|---|---|---|---|---|---|---|---|---|
| | **Population; setting** | **Prediction tools; PI classification system** | **Study design** | **Databases searched** | **Publication restrictions** (year; language; publication type) | **Quality assessment tool** | **Meta-analysis included; method of meta-analysis** | **N relevant studies in review** (n participants) | **N tools included** |
| Zimmerman[109] (2018)<br><br>Accuracy | Adult inpatients; ICU | Any scale or index; NS | NS | MEDLINE; CINAHL; COCHRANE; El Banco de Datos de Enfermería; nursing database; LILACS | 1962-2016; English, Portuguese, Spanish; NS | None | No | 13 | 11 |

AHCPR – Agency for Health Care Policy and Research; CASP – Critical Appraisal Skills Programme; CNKI – China National Knowledge Infrastructure; COSMIN - COnsensus-based Standards for the selection of health Measurement INstruments; CUIDEN - Bibliographic Database Index Foundation including scientific production on Health Care in Latin American; DARE – Database of Abstracts of Reviews of Effects; DEV – model development study; EPUAP – European Pressure Ulcer Advisory Panel; FN – false negative; FP – false positive; ICU – intensive care unit; ICD-9 – International Classification of Diseases Ninth Edition; JBI – Joanna Briggs Institute; LILACS – Latin America and Caribbean Health Sciences Literature; ML – machine learning; NOS – Newcastle Ottawa Scale; NS – not stated; NPUAP – National Pressure Ulcer Advisory Panel; OR – odds ratio; PI – pressure injury; PROBAST – Prediction model Risk of Bias Assessment; QUADAS – Quality Assessment of Diagnostic Accuracy Studies; RoB – risk of bias; (S)ROC – (summary) receiver operating curve; TN - true negative; TP – true positive; ULAKBIM – Turkish Academic Network and Information Center; VAL – model validation study

[a]Patients and HCWs

[b]KQ1 – key question 1 looks at effectiveness of risk assessment tools; KQ2 – key question 2 looks at diagnostic accuracy/validity of risk assessment tools

[b]Version of modified Norton scale cannot be determined.

Table A3. AMSTAR-2 assessment results per review

| Review author (pub. year) | ITEM 1 | ITEM 2 | ITEM 3 | ITEM 4 | ITEM 5 | ITEM 6 | ITEM 7 | ITEM 8 | ITEM 9 | ITEM 10 | ITEM 11 | ITEM 12 | ITEM 13 | ITEM 14 | ITEM 15 | Overall confidence |
|---|---|---|---|---|---|---|---|---|---|---|---|---|---|---|---|---|
| **Prognostic accuracy reviews** | | | | | | | | | | | | | | | | |
| Chen[85] (2023) | N | N | N | PY | Y | Y | N | PY | Y | N | N | N | N | N | Y | **Critically Low** Y=3/15 PY=2/15 N=10/15 |
| Chen[86] (2016) | N | N | N | PY | Y | N | N | PY | PY | N | N | N | N | Y | Y | **Critically Low** Y=3/15 PY=3/15 N=9/15 |
| Chou[87] (2013) | Y | Y | N | Y | Y | N | Y | Y | PY | Y | N | Y | Y | N | Y | **Low** Y=10/15 PY=1/15 N=4/15 |
| Garcia-Fernandez[88] (2014) | N | N | Y | PY | N | N | N | N | N | N | Y | Y | Y | N | N | **Critically Low** Y=4/15 PY=1/15 N=10/15 |
| He[90] (2012) | N | N | N | N | Y | N | N | N | Y | N | N | N | N | Y | Y | **Critically Low** Y=4/15 PY=0/15 N=11/15 |
| Huang[92] (2021) | N | Y | N | PY | Y | Y | N | N | Y | N | Y | N | N | Y | Y | **Critically Low** Y=7/15 PY=1/15 N=7/15 |
| Mehicic[96] (2024) | N | Y | N | N | Y | Y | N | N | N | N | NA | NA | N | Y | Y | **Critically Low** Y=5/13 PY=0/13 N=8/13 |
| Pancorbo-Hidalgo[110] (2006) | N | N | Y | PY | N | Y | N | PY | N | N | N | Y | Y | N | N | **Critically Low** Y=4/15 PY=2/15 N=9/15 |
| Park[99] (2016a) | N | N | N | PY | N | N | N | PY | N | N | N | N | N | Y | Y | **Critically Low** Y=2/15 |

| **Review author** (pub. year) | **ITEM 1** | **ITEM 2** | **ITEM 3** | **ITEM 4** | **ITEM 5** | **ITEM 6** | **ITEM 7** | **ITEM 8** | **ITEM 9** | **ITEM 10** | **ITEM 11** | **ITEM 12** | **ITEM 13** | **ITEM 14** | **ITEM 15** | **Overall confidence** |
|---|---|---|---|---|---|---|---|---|---|---|---|---|---|---|---|---|
| | | | | | | | | | | | | | | | | PY=2/15<br>N=11/15 |
| Park[100] (2016b) | N | N | N | PY | N | Y | N | PY | N | N | N | N | N | Y | Y | **Critically Low**<br>Y=3/15<br>PY=2/15<br>N=10/15 |
| Park[101] (2015) | N | N | N | PY | Y | Y | N | PY | N | N | N | Y | Y | Y | Y | **Critically Low**<br>Y=6/15<br>PY=2/15<br>N=7/15 |
| Pei[102] (2023) | N | Y | N | Y | N | Y | N | N | Y | N | N | N | Y | N | Y | **Critically Low**<br>Y=6/15<br>PY=0/15<br>N=9/15 |
| Qu[103] (2022) | N | Y | N | N | Y | Y | N | N | Y | N | N | N | Y | N | Y | **Critically Low**<br>Y=6/15<br>PY=0/15<br>N=9/15 |
| Tayyib[104] (2013) | N | N | N | PY | N | N | N | N | N | N | NA | NA | N | N | N | **Critically Low**<br>Y=0/13<br>PY=1/13<br>N=12/13 |
| Wang[105] (2022) | N | N | N | PY | Y | Y | N | N | N | N | Y | N | Y | N | Y | **Critically Low**<br>Y=5/15<br>PY=1/15<br>N=9/15 |
| Wei[106] (2020) | N | N | N | PY | Y | Y | N | N | PY | N | N | N | Y | Y | N | **Critically Low**<br>Y=4/15<br>PY=2/15<br>N=9/15 |
| Wilchesky[107] (2015) | N | N | N | PY | N | N | N | N | N | N | N | N | N | Y | Y | **Critically Low**<br>Y=2/15<br>PY=1/15<br>N=12/15 |
| Zhang[108] (2021) | N | Y | N | PY | Y | Y | N | PY | Y | N | Y | N | Y | Y | Y | **Low**<br>Y=8/15 |

| Review author (pub. year) | ITEM 1 | ITEM 2 | ITEM 3 | ITEM 4 | ITEM 5 | ITEM 6 | ITEM 7 | ITEM 8 | ITEM 9 | ITEM 10 | ITEM 11 | ITEM 12 | ITEM 13 | ITEM 14 | ITEM 15 | Overall confidence |
|---|---|---|---|---|---|---|---|---|---|---|---|---|---|---|---|---|
| | | | | | | | | | | | | | | | | PY=2/15<br>N=5/15 |
| Zimmerman [109] (2018) | N | N | N | N | Y | N | N | N | N | N | NA | NA | N | N | N | **Critically Low**<br>Y=1/13<br>PY=0/13<br>N=12/13 |
| **Summary** | **1/19 Yes** | **6/19 Yes** | **2/19 Yes** | **2/19 Yes 13/19 PY** | **12/19 Yes** | **11/19 Yes** | **1/19 Yes** | **1/19 Yes 7/19 PY** | **6/19 Yes 3/19 PY** | **1/19 Yes** | **4/16 Yes** | **4/16 Yes** | **9/19 Yes** | **10/19 Yes** | **14/19 Yes** | |
| **Clinical effectiveness reviews** | | | | | | | | | | | | | | | | |
| Baris[84] (2015) | N | N | N | PY | N | Y | N | N | N | N | NA | NA | N | N | N | **Critically Low**<br>Y=1/13<br>PY=1/13<br>N=11/13 |
| Chou[87] (2013) | Y | Y | N | Y | Y | N | Y | Y | Y | Y | NA | NA | Y | N | Y | **Moderate**<br>Y=10/13<br>PY=0/13<br>N=3/13 |
| Gaspar[89] (2019) | Y | N | N | PY | Y | N | N | Y | N | N | NA | NA | N | Y | Y | **Critically Low**<br>Y=5/13<br>PY=1/13<br>N=7/13 |
| Health Quality Ontario[91] (2009) | N | N | N | N | N | N | N | Y | N | N | NA | NA | Y | N | Y | **Critically Low**<br>Y=3/13<br>PY=0/13<br>N=10/13 |
| Kottner[93] (2009) | N | N | N | PY | Y | Y | N | Y | PY | N | NA | NA | Y | Y | Y | **Critically Low**<br>Y=6/13<br>PY=2/13<br>N=5/13 |
| Lovegrove[94] (2021) | Y | PY | N | PY | Y | Y | N | Y | PY | N | NA | NA | Y | Y | Y | **Low**<br>Y=7/13<br>PY=3/13<br>N=2/13 |

| Review author (pub. year) | ITEM 1 | ITEM 2 | ITEM 3 | ITEM 4 | ITEM 5 | ITEM 6 | ITEM 7 | ITEM 8 | ITEM 9 | ITEM 10 | ITEM 11 | ITEM 12 | ITEM 13 | ITEM 14 | ITEM 15 | Overall confidence |
|---|---|---|---|---|---|---|---|---|---|---|---|---|---|---|---|---|
| Lovegrove[95] (2018) | N | Y | N | PY | Y | Y | N | Y | PY | N | NA | NA | Y | Y | Y | **Low** Y=7/13 PY=2/13 N=4/13 |
| Mehicic[96] (2024) | Y | Y | N | N | Y | Y | N | N | PY | N | NA | NA | Y | N | Y | **Critically Low** Y=6/13 PY=1/13 N=6/13 |
| Moore[97] (2019) | Y | Y | N | Y | Y | N | Y | Y | Y | Y | NA | NA | Y | Y | Y | **High** Y=11/13 PY=0/13 N=2/13 |
| Pancorbo-Hidalgo[110] (2006) | N | N | Y | PY | N | Y | N | Y | PY | N | NA | NA | Y | N | N | **Critically Low** Y=4/13 PY=3/13 N=6/13 |
| Tayyib[104] (2013) | N | N | N | PY | N | N | N | PY | N | N | NA | NA | N | N | N | **Critically Low** Y=0/13 PY=2/13 N=11/13 |
| **Summary** | **4/11 Yes** | **4/11 Yes 1/11 PY** | **1/11 Yes** | **2/11 Yes 7/11 PY** | **7/11 Yes** | **6/11 Yes** | **2/11 Yes** | **8/11 Yes 1/11 PY** | **2/11 Yes 5/11 PY** | **2/11 Yes** | **11/11 NA** | **11/11 NA** | **8/11 Yes** | **5/11 Yes** | **8/11 Yes** | |

Item 1 – Adequate research question/ inclusion criteria?; Item 2 – Protocol and justifications for deviations?; Item 3 – Reasons for study design inclusions?; Item 4 – Comprehensive search strategy?; Item 5 – Study selection in duplicate?; Item 6 – Data extraction in duplicate?; Item 7 – Excluded studies list (with justifications)?; Item 8 – Included studies description adequate?; Item 9 – Assessment of RoB/quality satisfactory?; Item 10 – Studies' sources of funding reported?; Item 11 – Appropriate statistical synthesis method?; Item 12 – Assessment of impact of RoB on synthesised results?; Item 13 – Assessment of impact of RoB on review results?; Item 14 – Discussion/investigation of heterogeneity?; Item 15 – Conflicts of interest reported?
RoB – Risk of Bias; Y – Yes; PY – Partial Yes; N – No. Further details on AMSTAR items are given in Appendix 4.

#### Table A4. Statistical synthesis results of accuracy (from all 16 reviews, that conducted meta-analysis or statistical synthesis), by prediction tool

| **Review author** (publication year) | **n studies; N partic-ipants** | **Brief description of included studies** | **Brief description of included study quality** | **Summary estimates of accuracy parameters (main results from statistical syntheses)** | | | | | |
|---|---|---|---|---|---|---|---|---|---|
| | | | | **Sensitivity (95% CI)** | **Specificity (95% CI)** | **Likelihood ratios (95% CI)** | **DOR (95% CI)** | **AUROC (95% CI)** | **Predictive values (95% CI)** |
| **TOOL: Braden (1987)** [111 112] | | | | | | | | | |
| Huang[92] (2021) | n = 60;<br>N = 49,326 | Setting: hospital (n=45; includes 22 in ICU or other acute units), LTCF (n=15)<br>Sample size: 25 to 10,098<br>Mean age: range 31.7±10.9 to 84.6±7.9<br>Design: 47 prospective, 13 retrospective<br>Braden cut-off (out of 23): range ≤10 to ≤20 | QUADAS-II:<br>Patient selection: low RoB in 11 (18%) studies, unclear RoB (>50%); low concern about applicability in 44 (73%) studies, high concern for applicability in remaining 16;<br><br>Index test: low RoB in 39 (65%) studies, high RoB (approx. 33%); low concern about applicability in 51 (85%) studies;<br><br>Reference standard: low RoB in 58 (97%) studies, unclear RoB in 2 (3%); low concern about applicability in 51 (85%) studies, unclear concern in 9 (15%);<br><br>Flow and timing: low RoB in 50 (83%) studies; unclear RoB in remaining 10 (17%). | **0.78 (0.74, 0.82)**<br>N=15,241<br><br>By cut-off:<br><br>*≤15* (n=15): 0.79 (0.76, 0.82)<br><br>*16* (n=19): 0.75 (0.67, 0.82)<br><br>*17* (n=4): 0.69 (0.61, 0.76)<br><br>*18* (n=15): 0.82 (0.73, 0.89)<br><br>*≥19* (n=7): 0.78 (0.65, 0.87) | **0.72 (0.66, 0.78)**<br>N=34,085<br><br>By cut-off:<br><br>*≤15*:<br>0.66 (0.55, 0.75)<br><br>*16*:<br>0.85 (0.70, 0.93)<br><br>*17*:<br>0.86 (0.50, 0.97)<br><br>*18*:<br>0.70 (0.62, 0.77)<br><br>*≥19*:<br>0.54 (0.44, 0.63) | **PLR 2.80 (2.30, 3.50)**<br><br>**NLR 0.30 (0.26, 0.35)** | **9.00 (7.00, 13.00)**<br><br>By cut-off:<br><br>*≤15*:<br>7.00 (4.00, 12.00)<br><br>*16*:<br>17.00 (8.00, 36.00)<br><br>*17*:<br>14.00 (2.00, 103.00)<br><br>*18*:<br>11.00 (6.00, 20.00)<br><br>*≥19*:<br>4.00 (2.00, 7.00) | **0.82 (0.79, 0.85)**<br><br>By cut-off:<br><br>*≤15*:<br>0.80 (0.76, 0.83)<br><br>*16*:<br>0.84 (0.80, 0.87)<br><br>*17*:<br>0.73 (0.69, 0.77)<br><br>*18*:<br>0.83 (0.79, 0.86)<br><br>*≥19*:<br>0.67 (0.63, 0.71) | |
| Zhang[108] (2021) | n = 18;<br>N = 11,167 | Overview of studies on Braden not reported.<br><br>Braden cut-off points used range from 10.5 to 20. | Overview of studies on Braden not reported.<br><br>QUADAS-II:<br>Overall judgement was "not so satisfactory". | **0.78 (0.68, 0.85)** | **0.61 (0.40, 0.79)** | **PLR 2.00 (1.24, 3.24)**<br><br>**NLR 0.36 (0.25, 0.52)** | **5.52 (2.61, 11.67)** | **0.78** | |

| Review author (publication year) | n studies; N partic-ipants | Brief description of included studies | Brief description of included study quality | Summary estimates of accuracy parameters (main results from statistical syntheses) | | | | | |
|---|---|---|---|---|---|---|---|---|---|
| | | | | Sensitivity (95% CI) | Specificity (95% CI) | Likelihood ratios (95% CI) | DOR (95% CI) | AUROC (95% CI) | Predictive values (95% CI) |
| Wei[106] (2020) | n = 11; N = 10,044 | Prospective (7/11); retrospective (4/11). ICU settings. Mean age from 49.2±17.3 to 62.5±16.3. Classification systems: PPPU (n=1), NPUAP(n=5), EPUAP (n=2), ICD-9 (n=1), NS (n=2). Braden scale cut-off: 11 (n=2), 12 (n=1), 13 (n=3), 14 (n=2), 16 (n=2), 18 (n=1) | QUADAS-II: Limited detail. No study was considered low risk and low concern for applicability in all domains.<br><br>All 11 studies met 80% "low risk" or "yes" in 13 questions. | **0.89 (0.87, 0.91)**<br><br>n=1123<br>$I^2$ = 94.9%, p<0.001<br><br>Q*=0.720 (0.66, 0.78) (SE 0.03) | **0.28 (0.27, 0.29)**<br><br>n=8921<br>$I^2$ = 99.2%, p<0.001 | | **6.29 (4.09, 9.68)** | **0.78 (0.72, 0.85)** | |
| Chen[86] (2016) | n = 8; N = 41,489 | Settings: nursing homes, LTCF, tertiary care hospitals, veterans' medical centres, and skilled nursing facilities. NPUAP used as reference standard, Stage I-IV (prevalence 6.4% to 30.1%). Braden scale cut-off ranged from 17 to 20. | QUADAS (original): All studies scored 'No' for two items (reporting of uninterpretable/intermediate results and explanation of any withdrawals). Five studies judged 'Yes' on remaining 9 QUADAS items; 3 studies judged 'Unclear' for reporting of selection criteria. | **0.80 (0.79, 0.81)**<br><br>$I^2$ = 97.4%; Q* 0.709 (0.63, 0.79) (SE 0.04) | **0.42 (0. 42, 0.43)**<br><br>$I^2$ = 98.7% | | **5.66 (3.77, 8.48)**<br><br>$I^2$ = 96.4%; $\tau^2$ = 0.4173 | **0.769 (0.675, 0.862)**<br><br>(SE 0.048) | |
| Park[100] (2016b) | n = 25; N = 10,547 | Overview of studies on Braden not reported. Cut-offs selected "by following the one which the study researcher(s) indicated to be the most effective". Braden cut-off: 13 (n=2); 16 (n=8); 17 (n=2); 18 (n=9); 19 (n=3); 20 (n=1) | Overview of studies on Braden not reported. | **0.72 (0.69, 0.74)**<br><br>$I^2$ = 79.9% ($\chi^2$=119.57, p<0.001)<br><br>Q* = 0.72 (SE = 0.02) | **0.63 (0.62, 0.64)**<br><br>$I^2$ = 96.4% ($\chi^2$=673.34, p<0.001) | **PLR 2.31 (1.98, 2.69)**<br><br>**NLR 0.43 (0.36, 0.51)** | **6.50 (4.64, 9.11)** | **0.79**<br>(SE = 0.02) | |
| Park[101] (2015) | n = 21; N = 6,070 | Settings: ICU, hospital, nursing units, trauma centre Average age: >50% of studies in 50s or 60s<br>Reference standard: NPUAP | QUADAS-II: "None of the studies were evaluated to have a high risk of bias in each area. ... 1) they only included prospective studies, 2) | **0.72 (0.68, 0.75)**<br><br>$I^2$ = 64.0% ($\chi 2$ = 55.48, p<0.001) | **0.81 (0.80, 0.82)**<br><br>$I^2$ = 96.2% ($\chi 2$ = 25.38, p<0.001) | **PLR 3.43 (2.66, 4.44)**<br><br>**NLR 0.38 (0.30, 0.48)** | **10.30 (6.65, 15.96)** | **0.84**<br>(SE 0.02) | |

| Review author (publication year) | n studies; N partic-ipants | Brief description of included studies | Brief description of included study quality | Summary estimates of accuracy parameters (main results from statistical syntheses) | | | | | |
|---|---|---|---|---|---|---|---|---|---|
| | | | | Sensitivity (95% CI) | Specificity (95% CI) | Likelihood ratios (95% CI) | DOR (95% CI) | AUROC (95% CI) | Predictive values (95% CI) |
| | | (n=10), AHCPR (n=2), TDCPS (n=1), tools developed by individual researchers (n=6); NS (n=2).<br>Evaluation conducted at time of or within 24-72hr of hospitalization. Cut-off points ranged from <13 to <20. | test diagnosis measure and reference standards test were non-invasive and evaluated by regular observation by nurses, which means that almost none of the patients were excluded." "All selected studies were confirmed to be of high quality and meet all areas of the quality evaluation" | | | | | | |
| Wilchesky [107] (2015) | n = 11;<br>N = 40,361 | Assessed predictive validity (n=4; 1,145 patients); mean PI prevalence 20.71%; Braden cut-off 18<br><br>Assessed concurrent/ screening validity (n=7; 39,216 patients); mean PI prevalence 15.85%; Braden cut-off 20 in all except one study using 18 | Not done | **0.86 (0.85, 0.87)**<br><br>Overall: mean 0.84 (SD = 0.083, range 0.70–0.95)<br><br>Predictive: mean 0.74<br>Concurrent: mean 0.89 | **0.39 (0.38, 0.39)**<br><br>Overall: mean 0.51 (SD = 0.168, range 0.34–0.74)<br><br>Predictive: mean 0.72<br>Concurrent: mean 0.39 | | RR reported.<br>**RR = 4.33 (3.28, 5.72)**<br>χ2=107.51, p<0.001<br><br>Predictive: RR = 4.53 (3.42, 6.00)<br>χ2=111.65, p<0.001<br>Concurrent: RR = 4.22 (3.02, 5.89)<br>χ2 = 71.2, p<0.001 | | Overall:<br>**mean PPV 0.28**<br>(SD=0.151, range 0.09–0.54);<br>**mean NPV 0.93**<br>(SD=0.053, range 0.82–0.98)<br><br>Predictive validity: mean PPV 0.42, NPV 0.91<br><br>Concurrent validity: mean PPV 0.21, NPV 0.94 |

| Review author (publication year) | n studies; N partic-ipants | Brief description of included studies | Brief description of included study quality | Summary estimates of accuracy parameters (main results from statistical syntheses) | | | | | |
|---|---|---|---|---|---|---|---|---|---|
| | | | | **Sensitivity (95% CI)** | **Specificity (95% CI)** | **Likelihood ratios (95% CI)** | **DOR (95% CI)** | **AUROC (95% CI)** | **Predictive values (95% CI)** |
| Garcia-Fernandez [88] (2014) | n = 33; N = 8,615 | Not reported | Not reported.<br>Included studies passed CASP quality assessment.<br><br>(13/86 studies excluded across whole review) | | | | RR reported.<br>**RR = 4.26 (3.27, 5.55)**<br><br>$I^2$ = 68%;<br>$\tau^2$ = 0.37 | | |
| Chou[87] (2013) | n = 32 (33 publica-tions);<br>N = 11,596 | Overview of studies on Braden not reported.<br><br>Braden cut-offs ranged between ≤10 and ≤20. The majority of studies used cut-offs of ≤15 (n=12), ≤16/<17 (n=11) or ≤18 (n=16). | Overview of studies on Braden not reported.<br><br>Criteria based on QUADAS-II: Studies evaluating Braden: Rated good quality (n=13), fair quality (n=18) and poor quality (n=2). | Median[A] by cut-off:<br><br>*≤10* (n=1): 0.91<br><br>*≤15* (n=12): 0.33 (range 0.09 - 0.82)<br><br>*≤16* (n=8): 0.77 (range 0.35 - 1.0)<br><br>*≤18* (n=16): 0.74 (range 0.33 - 1.0)<br><br>*≤20* (n=1): 0.97 | Median[A] by cut-off:<br><br>*≤10* (n=1): 0.96<br><br>*≤15* (n=12): 0.91 (range 0.67 - 0.95)<br><br>*≤16* (n=8): 0.64 (range 0.14 - 1.0)<br><br>*≤18* (n=16): 0.68 (range 0.34 - 0.86)<br><br>*≤20* (n=1): 0.05 | PLR ranged from 22.75 (cut-off ≤10) and 1.00 (cut-off <17).<br><br>NLR ranged from 0.09 (cut-off ≤10) and 1.00 (cut-off <17). | | Median in n=7 studies that reported AUROC.<br><br>**0.77 (range 0.55 - 0.88)** | |
| He[90] (2012) | n = 3;<br>N = 609 | Study countries: South Korea, Germany and USA; Study design: two prospective, one NS; all surgical ICU populations; mean age ranges from 58.1-62.0 years; Assessment time: two pre-operative, one NS; | QUADAS (original):<br>Two studies at high risk of bias due to the spectrum of patients not being representative of those who will receive the test in practice. One study at unclear risk of bias. It was unclear whether the whole sample or a random sample | **0.42 (0.38, 0.47)** | **0.84 (0.83, 0.85)** | | **4.40 (2.98, 6.50)** | **0.6921** (SE 0.0346) | |

| Review author (publication year) | n studies; N partic-ipants | Brief description of included studies | Brief description of included study quality | Summary estimates of accuracy parameters (main results from statistical syntheses) | | | | | |
|---|---|---|---|---|---|---|---|---|---|
| | | | | **Sensitivity (95% CI)** | **Specificity (95% CI)** | **Likelihood ratios (95% CI)** | **DOR (95% CI)** | **AUROC (95% CI)** | **Predictive values (95% CI)** |
| | | prevalence: 18.3%, 49.0%, 4.7%; Cut off points: one study used 14, two studies used various ranging from 9-20, and 5-23. | received a diagnosis with the reference standard and withdrawals from the study were unclear. | | | | | | |
| Pancorbo-Hildago[110] (2006) | n = 20;<br>N = 6,443 (included in aggregated analysis)<br><br>n = 16;<br>N = 5,847 (included in MA) | Hospital medical, cardiovascular, orthopaedic and surgical units, hospital acute care, hospital extended care, home care, hospice, long-term care facilities, veterans administration medical centres, skilled nursing facilities, hospital internal medicine, hospital cardiac surgery; classification system not reported but minimum PI stage I or II; Braden cut-offs ≤14, 16, 17, 18, 19, 20; follow-up period ranges from 5 days to 3 months/whole stay in unit/discharge/ death | Studies excluded if considered not to be 'valid'. This was determined by assessment of methodological quality; CASP guide for RCTs and a clinical assessment guide developed for the clinical practice guide for PI assessment and prevention for prospective cohort studies. | **0.57** | **0.68** | | **OR = 4.08 (2.56, 6.48)** | | **PPV 0.23**<br>**NPV 0.91** |
| **TOOL: Modified Braden scales: Braden – modified by Song & Choi[113] (1991)** | | | | | | | | | |
| Park[99] (2016a) | n = 4;<br>N = 688 | Prospective (4/4), recruiting patients with no PI at baseline (hospital ward (n=2) or ICU (n=3); mean age in the 50s (n=2), 60s (n=2). Classification used: AHCPR (n=3), Bergstrom (n=1). Braden scale cut-off used: <21 (n=1), <23 (n=1), <24 (n=2) | QUADAS-II:<br>"None had 'high risk'" | **0.97 (0.92, 0.99)**<br><br>n=125<br>Q* 0.90 (0.03) | **0.70 (0.66, 0.73)**<br><br>n=563<br>Q* 0.90 (0.03) | **PLR 3.47 (1.33, 9.06)**<br><br>**NLR 0.08 (0.04, 0.19)** | **56.56 (21.88, 146.21)** | **0.95** (SE 0.02) | |

| Review author (publication year) | n studies; N partic-ipants | Brief description of included studies | Brief description of included study quality | Summary estimates of accuracy parameters (main results from statistical syntheses) | | | | | |
|---|---|---|---|---|---|---|---|---|---|
| | | | | Sensitivity (95% CI) | Specificity (95% CI) | Likelihood ratios (95% CI) | DOR (95% CI) | AUROC (95% CI) | Predictive values (95% CI) |
| Garcia-Fernandez [88] (2014) | n = 3;<br>N = 476 | Not reported | Not reported.<br>Included studies passed CASP quality assessment. | | | | RR reported.<br>**RR = 26.06 (9.01, 75.39)**<br><br>$I^2$ = 0%;<br>$\tau^2$ = 0 | | |
| **TOOL: Braden – modified by Pang & Wong[114] (1998)** | | | | | | | | | |
| Park[99] (2016a) | n = 2;<br>N = 626 | Prospective (2/2), recruiting patients with no PI at baseline (OS ward (n=1) or NS (n=1); mean age 79.4 and 54.1.<br>Classification used: NPUAP (n=2)<br>Braden scale cut-off used: <19 (n=1), <14 (n=1) | QUADAS-II:<br>"None had 'high risk'" | **0.89 (0.71, 0.98)**<br><br>n=27<br>Q* not calculated | **0.71 (0.67, 0.75)**<br><br>n=599<br>Q* not calculated | **PLR 2.87 (1.88, 4.38)**<br><br>**NLR 0.17 (0.06, 0.49)** | **16.06 (4.75, 54.35)** | Not calculated | |
| **TOOL: Braden – modified by Kwong[115] (2005)** | | | | | | | | | |
| Garcia-Fernandez [88] (2014) | n = 2;<br>N = 626 | Not reported | Not reported.<br>Included studies passed CASP quality assessment. | | | | RR reported.<br>**RR = 13.68 (4.19, 44.64)**<br><br>$I^2$ = 0%;<br>$\tau^2$ = 0 | | |
| **TOOL: Cubbin & Jackson[116] (1991)** | | | | | | | | | |
| Chen[85] (2023) | n = 9;<br>N = 7,684 | Study designs included prospective studies (n=2), a prospective and cross-sectional study (n=1), retrospective studies (n=3), an observational study (n=1), a predictive correlational study (n=1) and a longitudinal study (n=1). Studies were conducted in | Authors state that *"none of [the 9 included studies] has a high risk of bias in any field"*, despite indicating some as 'high risk' in the table of QUADAS-2 results.<br><br>QUADAS-2:<br>Patient selection: low RoB in 5 (56%) studies, unclear RoB in 1 (11%) study, high RoB in 3 (33%), | **0.81 (0.51, 0.95)**<br><br>n=1,558 | **0.76 (0.58, 0.88)**<br><br>n=6,126 | **PLR 3.34 (2.14, 5.21)**<br><br>**NLR 0.25 (0.09, 0.68)** | **13.24 (5.41, 32.40)** | **0.84 (0.81, 0.87)** | |

| Review author (publication year) | n studies; N partic-ipants | Brief description of included studies | Brief description of included study quality | Summary estimates of accuracy parameters (main results from statistical syntheses) | | | | | |
|---|---|---|---|---|---|---|---|---|---|
| | | | | **Sensitivity (95% CI)** | **Specificity (95% CI)** | **Likelihood ratios (95% CI)** | **DOR (95% CI)** | **AUROC (95% CI)** | **Predictive values (95% CI)** |
| | | South Korea (n=3), the US (n=2), Turkey (n=2), China (n=1) and Portugal (n=1). Studies targeted research participants >18 years old (n=6), >16 years old (n=1), ≥21 years old (n=1) or did not set an age limit (n=1). All studies restricted to patients without PIs on ICU admission. | low applicability concerns in all studies;<br>Index test: low RoB in 4 (44%) studies, unclear RoB in 3 (33%) studies, high RoB in 2 (22%) studies, low applicability concerns in 7 (78%) studies, high applicability concerns in 2 (22%) studies;<br>Reference standard: low RoB in 1 (11%) study, unclear RoB in 6 (67%) studies), high RoB in 2 (22%) studies, low applicability concerns in 4 (44%) studies, unclear applicability concerns in 2 (22%) studies, high applicability concerns in 3 (33%) studies;<br>Flow and timing: low RoB in 7 (78%) studies, unclear RoB in 2 (22%) studies | | | | | | |
| Zhang[108] (2021) | n = 6;<br>N = 800 | Overview of studies on C&J not reported.<br><br>All 6 prospective studies. | Overview of studies on C&J not reported. | **0.84 (0.59, 0.95)** | **0.84 (0.66, 0.93)** | **PLR 5.12 (2.70, 9.70)**<br><br>**NLR 0.19 (0.08, 0.49)** | **26.45 (13.51, 51.78)** | **0.9** | |
| Park[99] (2016a) | n = 4;<br>N = 662 | Prospective (4/4); ICU patients for all studies (1 in surgical ICU), with no PI at baseline (n=3); mean age in the 50s (n=2), 60s (n=2). Classification used: AHCPR (n=2), NPUAP (n=1), Lowthian (n=1). | QUADAS-II:<br>"None had 'high risk'" | **0.67 (0.60, 0.74)**<br><br>n=194<br>Q* = 0.75 (0.06) | **0.75 (0.71, 0.79)**<br><br>n=468. Q* 0.75 (0.06) | **PLR 2.80 (1.66, 4.72)**<br><br>**NLR 0.34 (0.15, 0.76)** | **9.46 (2.41, 37.22)** | **0.82**<br>(SE 0.06) | |

| Review author (publication year) | n studies; N partic-ipants | Brief description of included studies | Brief description of included study quality | Summary estimates of accuracy parameters (main results from statistical syntheses) | | | | | |
|---|---|---|---|---|---|---|---|---|---|
| | | | | **Sensitivity (95% CI)** | **Specificity (95% CI)** | **Likelihood ratios (95% CI)** | **DOR (95% CI)** | **AUROC (95% CI)** | **Predictive values (95% CI)** |
| | | C&J scale cut-off used: <24 (n=2), <26 (n=1), <28 (n=1) | | | | | | | |
| Garcia-Fernandez [88] (2014) | n = 3; N = 370 | Not reported | Overview of studies on C&J not reported. | | | | RR reported. **RR = 8.63 (3.02, 24.66)** $I^2$ = 65%; $\tau^2$ = 0.55 | | |
| **TOOL: Cubbin & Jackson – Revised: "Jackson & Cubbin"[117] (1999)** | | | | | | | | | |
| Garcia-Fernandez [88] (2014) | n = 2; N = 259 | Not reported | Overview of studies on J&C not reported. | | | | RR reported. **RR = 3.16 (1.49, 6.71)** $I^2$ = 0%; $\tau^2$ = 0 | | |
| **TOOL: EMINA[118] (2001)** | | | | | | | | | |
| Garcia-Fernandez [88] (2014) | n = 2; N = 861 | Not reported | Overview of studies on EMINA not reported. | | | | RR reported. **RR=6.17 (3.46, 11.01)** $I^2$ = 0%; $\tau^2$ = 0 | | |
| **TOOL: EVARUCI[119] (2001)** | | | | | | | | | |
| Zhang[108] (2021) | n = 3; N = 3,063 | Overview of studies on EVARUCI not reported. All 3 prospective studies. | Overview of studies on EVARUCI not reported. | **0.84 (0.79, 0.89)** | **0.68 (0.66, 0.70)** | **PLR 2.32 (2.14, 2.51)** **NLR 0.25 (0.19, 0.35)** | **9.79 (6.81, 14.07)** | **0.82** | |
| **TOOL: Norton[120] (1962)** | | | | | | | | | |
| Park[99] (2016a) | n = 7; N = 2,899 | Prospective (6/7); inpatients with no PI at baseline (1 LTC, 2 'hospital', 1 ICU, 1 ICU & wards); mean age in the 50s (n=1), 60s (n=3), or 80s (n=1), | QUADAS-II: "None had 'high risk'" | **0.75 (0.70, 0.79)** n=383 | **0.57 (0.55, 0.59)** n=2516 | **PLR 1.77 (1.26, 2.50)** **NLR 0.49 (0.32-0.76)** | **7.57 (2.53, 22.64)** | **0.82** (SE 0.05) | |

| **Review author** (publication year) | **n studies; N partic-ipants** | **Brief description of included studies** | **Brief description of included study quality** | **Summary estimates of accuracy parameters (main results from statistical syntheses)** | | | | | |
|---|---|---|---|---|---|---|---|---|---|
| | | | | **Sensitivity (95% CI)** | **Specificity (95% CI)** | **Likelihood ratios (95% CI)** | **DOR (95% CI)** | **AUROC (95% CI)** | **Predictive values (95% CI)** |
| | | or NS (n=2).<br>Classification used: AHCPR (n=3), NPUAP (n=2), EPUAP (n=1), TDCPS (n=1).<br>Nortcon scale cut-off used: <14 (n=2, but reported as 3 in paper), <15 (n=2), <16 (n=3) | | Q* 0.75 (SE=0.04) | Q* 0.75 (SE=0.04) | | | | |
| Park[100] (2016b) | n = 5;<br>N = 2,408 | Only reported overall, not by scale.<br>Norton cut-offs: 14 (n=2); 16 (n=3) | Only reported overall, not by scale. | **0.76 (0.71, 0.80)**<br><br>$I^2$ = 90.5% ($\chi^2$ = 41.97, p<0.001) | **0.55 (0.53, 0.57)**<br><br>$I^2$ 98.7% ($\chi^2$ =308.41, p<0.001) | **PLR 1.58 (1.07, 2.34)**<br><br>**NLR 0.47 (0.29, 0.76)** | **6.41 (1.72, 23.88)** | **0.84**<br>(SE 0.07) | |
| Garcia-Fernandez[88] (2014) | n = 16;<br>N = 5,032 | Not reported | Not reported.<br>Included studies passed CASP quality assessment. | | | | RR reported.<br>**RR = 3.69 (2.64, 5.16)**<br><br>$I^2$ = 66%;<br>$\tau^2$ = 0.23 | | |
| Chou[87] (2013) | n = 9;<br>N = 5,444 | Overview of studies on Norton not reported.<br><br>Norton cut-offs ranged between <12 and ≤16. The majority of studies used cut-offs of ≤14 (n=5), ≤16 (n=3). | Overview of studies on Norton not reported.<br><br>Criteria based on QUADAS-II: Rated as fair quality (n=6) and good quality (n=3). | Median[A] by cut-off:<br><br>*≤12* (n=1): 0.62<br><br>*≤14* (n=5): 0.75 (range 0.0 - 0.89)<br><br>*≤16* (n=3): 0, 0.75, 0.89 | Median[A] by cut-off:<br><br>*≤12* (n=1): 0.72<br><br>*≤14* (n=5): 0.68 (range 0.59 - 0.95)<br><br>*≤16* (n=3): 0.55, 0.59, 0.6. | PLR ranged from 1.83 (cut-off ≤16) to 2.34 (cut-off ≤14).<br><br>NLR ranged from 0.37 (cut-off ≤14) to 0.53 (cut-off <12). | | Median in n=3 studies that reported AUROC.<br><br>**0.74 (range 0.56 - 0.75)** | |

| Review author (publication year) | n studies; N partic-ipants | Brief description of included studies | Brief description of included study quality | Summary estimates of accuracy parameters (main results from statistical syntheses) | | | | | |
|---|---|---|---|---|---|---|---|---|---|
| | | | | **Sensitivity (95% CI)** | **Specificity (95% CI)** | **Likelihood ratios (95% CI)** | **DOR (95% CI)** | **AUROC (95% CI)** | **Predictive values (95% CI)** |
| Pancorbo-Hidalgo[110] (2006) | n = 5; N = 2,008 (included in aggregated analysis & MA) | Hospital cardiovascular surgery and neurosurgery, hospital orthopaedic surgery, geriatric centre, rehabilitation hospital medical and orthopaedic units, hospital medical units, surgical units and ICU; classification system not reported but minimum PI stage I (n=5) or II (n=1); Norton cut-offs ≤14 (n=2), ≤16 (n=3); follow-up period from 2 weeks-12 weeks/to discharge/death; average age, years = 53.1, 60.1, 80.4, NS (n=2). | All studies considered to be 'valid' through use of CASP. | **0.47** | **0.62** | | **OR = 2.16 (1.03, 4.54)** | | **PPV 0.18** **NPV 0.87** |
| **TOOL: Norton – modified by Ek[121] (1987)** | | | | | | | | | |
| Garcia-Fernandez [88] (2014) | n = 3; N = 502 | Not reported | Not reported. Included studies passed CASP quality assessment. | | | | RR reported. **RR = 2.38 (0.92, 6.12)** $I^2$ = 81%; $\tau^2$ = 0.56 | | |
| **TOOL: Norton – modified by Bienstein[122] (1991)** | | | | | | | | | |
| Garcia-Fernandez [88] (2014) | n = 2; N = 164 | Not reported | Not reported. Included studies passed CASP quality assessment. | | | | RR reported. **RR = 1.53 (1.11, 2.12)** $I^2$ =0%; $\tau^2$ =0 | | |

| Review author (publication year) | n studies; N partic-ipants | Brief description of included studies | Brief description of included study quality | Summary estimates of accuracy parameters (main results from statistical syntheses) | | | | | |
|---|---|---|---|---|---|---|---|---|---|
| | | | | Sensitivity (95% CI) | Specificity (95% CI) | Likelihood ratios (95% CI) | DOR (95% CI) | AUROC (95% CI) | Predictive values (95% CI) |
| **TOOL: PSPS[123] (1987)** | | | | | | | | | |
| Garcia-Fernandez [88] (2014) | n = 2;<br>N = 1,956 | Not reported | Not reported.<br>Included studies passed CASP quality assessment. | | | | RR reported.<br>**RR = 21.40 (10.74, 42.63)**<br><br>$I^2$ =0%;<br>$\tau^2$ = 0 | | |
| **TOOL: Waterlow[124] (1985)** | | | | | | | | | |
| Zhang[108] (2021) | n = 4;<br>N = 1,000 | Reported overall, not by scale.<br>All 4 prospective studies. | Reported overall, not by scale. | **0.63 (0.48, 0.76)** | **0.46 (0.22, 0.71)** | **PLR 1.16 (0.66, 2.01)**<br><br>**NLR 0.82 (0.40, 1.67)** | **1.42 (0.40, 5.07)** | **0.56** | |
| Park[99] (2016a) | n = 6;<br>N = 1,268 | Prospective (6/6); all male* inpatients aged over 60 on average with no PI at baseline (3 included ICU patients).<br>Classification used: AHCPR (n=2), NPUAP (n=2), EPUAP (n=1), TDCPS (n=1).<br>Waterlow scale cut-off used: <9 (n=1), <15 (n=1), <16 (n=2), <17 (n=1), NS (n=1)<br>* as reported in review's text. However, the table reports a mixture of female and male participants for all studies, with a mean female proportion of 50.73%. | QUADAS-II:<br>"None had 'high risk'" | **0.55 (0.49, 0.62)**<br><br>n=246<br>Q* 0.75<br>(SE=0.03) | **0.82 (0.80, 0.85)**<br><br>n=1222<br>Q* 0.75<br>(SE=0.03) | **PLR 2.89 (1.74, 4.79)**<br><br>**NLR 0.46 (0.31, 0.70)** | **9.22 (6.43, 13.23)** | **0.82**<br>(SE 0.03) | |
| Park[100] (2016b) | n = 5;<br>N = 1,406 | Only reported overall, not by scale. | Only reported overall, not by scale. | **0.53 (0.47, 0.60)** | **0.84 (0.81, 0.86)** | **PLR 3.09 (1.63, 5.83)** | **9.06 (6.30, 13.04)** | **0.81**<br>(SE 0.03) | |

| Review author (publication year) | n studies; N partic-ipants | Brief description of included studies | Brief description of included study quality | Summary estimates of accuracy parameters (main results from statistical syntheses) | | | | | |
|---|---|---|---|---|---|---|---|---|---|
| | | | | Sensitivity (95% CI) | Specificity (95% CI) | Likelihood ratios (95% CI) | DOR (95% CI) | AUROC (95% CI) | Predictive values (95% CI) |
| | | Waterlow cut-offs: 15 (n=1); 16 (n=2); 17 (n=1); NS (n=1) | | $I^2$ = 89.0% ($\chi^2$ = 36.31, p<0.001) | $I^2$ = 98.7% ($\chi^2$ =155.55, p<0.001) | **NLR 0.49 (0.34, 0.72)** | | | |
| Garcia-Fernandez [88] (2014) | n = 14; N = 3,969 | Not reported | Not reported. Included studies passed CASP quality assessment. | | | | RR reported. **RR = 2.66 (1.76, 4.01)** $I^2$ = 47%; $\tau^2$ = 0.19 | | |
| Chou[87] (2013) | n = 10; N = 3,905 | Overview of studies on Waterlow not reported. Waterlow cut-offs ranged from >9 to ≥20. | Overview of studies on Waterlow not reported. Criteria based on QUADAS-II: Studies evaluating Waterlow: Rated as good quality (n=2) and fair quality (n=8). | | | | | Median in n=4 studies that reported AUROC. **0.61 (range 0.54 - 0.66)** | |
| Pancorbo-Hidalgo[110] (2006) | n = 6; N = 2,246 (included in aggregated weighted mean analysis) n = 5; N = 2,215 (included in MA) | Hospital orthopaedic surgery, community, geriatric centre, rehabilitation hospital medical and orthopaedic units, hospital ICU, hospital medical, surgical and geriatric units; classification system not reported but minimum PI stage I (n=6) or II (n=1); Waterlow cut-offs ≥10 (n=4), NS (n=1), ≥16 (n=1), ≥15 (n=1); follow-up period from 2 weeks - 12 weeks / to discharge/death; average age, years in 50s (n=2), 60s (n=1), 80s (n=2), NS (n=2) | All studies considered to be 'valid' through use of CASP. | **0.82** | **0.27** | | **OR = 2.05 (1.11, 3.76)** | | **PPV 0.16 NPV 0.89** |

| Review author (publication year) | n studies; N partic-ipants | Brief description of included studies | Brief description of included study quality | Summary estimates of accuracy parameters (main results from statistical syntheses) | | | | | |
|---|---|---|---|---|---|---|---|---|---|
| | | | | Sensitivity (95% CI) | Specificity (95% CI) | Likelihood ratios (95% CI) | DOR (95% CI) | AUROC (95% CI) | Predictive values (95% CI) |
| **ML models:** | | | | | | | | | |
| Pei[102] (2023) | n = 14; N = 328,789 (included in MA) | Studies published from 2012-2022; studies conducted in China (n=4), Taiwan (n=3), USA (n=6), Germany (n=1), Japan (n=1), Australia (n=1), Spain (n=1) and Czech Republic (n=1) ;15 studies utilised retrospective data, while 3 used prospective data; studies focused on adult ICU inpatients (n=5), hospitalised patients (n=8), adult hospitalised patients awaiting surgery (n=3), cancer patients (n=1) and end-of-life adult inpatients (n=1); sample size range 168-149,006; number of patients with PI range 8-4663<br><br>Validation methods: both sample splitting and k-fold cross-validation (n= 10), sample splitting only (n=4), k-fold cross-validation only (n=1). External verification conducted (n=1). Validation methods not reported (n=2). Handling of missing data not disclosed (n=7). | PROBAST results:<br><br>Overall, 16/18 (88.9%) papers were at high ROB, 1 (5.6%) was at unclear ROB and only 1 (5.6%) was at low ROB.<br><br>14 (77.8%) studies were at high ROB in the analysis domain. The most common factors contributing to the high risk of bias in the analysis domain included an inadequate number of events per candidate predictor, poor handling of missing data and failure to deal with overfitting. | **0.79 (0.78, 0.80)**<br><br>n=18,807 | **0.87 (0.88, 0.87)**<br><br>n=309,982 | **PLR 10.71 (5.98, 19.19)**<br><br>**NLR 0.21 (0.08, 0.50)** | **52.39 (24.83, 110.55)** | **0.94** | |
| Qu[103] (2022) | n = 14; N = 118,292 | Only reported overall, not by algorithm type.<br>Conducted in: hospital | Only reported overall, not by algorithm type. | **0.66 (0.42, 0.84)** | **0.90 (0.78, 0.96)** | **PLR 6.9 (3.2, 14.7)** | **18 (7, 49)** | **0.88 (0.85, 0.91)** | |

| Review author (publication year) | n studies; N partic-ipants | Brief description of included studies | Brief description of included study quality | Summary estimates of accuracy parameters (main results from statistical syntheses) | | | | | |
|---|---|---|---|---|---|---|---|---|---|
| | | | | **Sensitivity (95% CI)** | **Specificity (95% CI)** | **Likelihood ratios (95% CI)** | **DOR (95% CI)** | **AUROC (95% CI)** | **Predictive values (95% CI)** |
| | | patients (n= 13); surgical patients (n=3), ICU (n=5), CVD patients (n=2), cancer patients (n=1), LTC (n=1)<br><br>**Decision Tree** models | QUADAS-II:<br>Five studies were judged high risk of bias for reference standard, 4 of which also had high concern for applicability in the same domain Five additional studies were judged unclear risk of bias in at least one domain.<br><br>Of those studies which included the DT algorithm, two studies had an unclear risk of bias and two studies were at high risk of bias. The rest were rated as low risk of bias. | n=7557 | n=110,735 | **NLR 0.37 (0.20, 0.69)** | | | |
| Qu[103] (2022) | n = 14;<br>N = 195,927 | **Logistic Regression** models | QUADAS-II:<br>Of those studies which included logistic regression, one study had an unclear risk of bias and four studies were at high risk of bias. The rest were rated as low risk of bias. | **0.71 (0.60, 0.80)**<br><br>n=9046 | **0.83 (0.75, 0.89)**<br><br>n=186,881 | **PLR 4.3 (3.1, 5.9)**<br><br>**NLR 0.35 (0.26, 0.46)** | **12 (9, 17)** | **0.84 (0.81, 0.87)** | |
| Qu[103] (2022) | n = 9;<br>N = 97,815 | **Neural Network** models | QUADAS-II:<br>Of those studies which included neural networks, one study had an unclear risk of bias and one study was at high risk of bias. The rest were rated as low risk of bias. | **0.73 (0.55, 0.86)**<br><br>n=9488 | **0.78 (0.65, 0.87)**<br><br>n=88,327 | **PLR 3.3 (2.1, 5.0)**<br><br>**NLR 0.35 (0.21, 0.59)** | **9 (5, 19)** | **0.82 (0.79, 0.85)** | |
| Qu[103] (2022) | n = 7;<br>N = 161,334 | **Random Forest** models | QUADAS-II:<br>Of those studies which included random forests, one study was at high risk of bias. The rest were rated as low risk of bias. | **0.72 (0.26, 0.95)**<br><br>n=5486 | **0.96 (0.80, 0.99)**<br><br>n=155,848 | **PLR 16.3 (2.4, 108.9)**<br><br>**NLR 0.29 (0.07, 1.29)** | **56 (3, 1258)** | **0.95 (0.93, 0.97)** | |

| Review author (publication year) | n studies; N partic-ipants | Brief description of included studies | Brief description of included study quality | Summary estimates of accuracy parameters (main results from statistical syntheses) | | | | | |
|---|---|---|---|---|---|---|---|---|---|
| | | | | **Sensitivity (95% CI)** | **Specificity (95% CI)** | **Likelihood ratios (95% CI)** | **DOR (95% CI)** | **AUROC (95% CI)** | **Predictive values (95% CI)** |
| Qu[103] (2022) | n = 9; N = 152,068 | **Support Vector Machine** models | QUADAS-II: Of those studies which included support vector machines, one study was at high risk of bias. The rest were rated as low risk of bias. | **0.81 (0.69, 0.90)** n=6562 | **0.81 (0.59, 0.93)** n=145,506 | **PLR 4.3 (1.8, 9.9)** **NLR 0.23 (0.13, 0.39)** | **19 (6, 54)** | **0.88 (0.85, 0.90)** | |

n – number of studies; N – number of participants; CI – confidence interval; DOR – diagnostic odds ratio; AUC/AUROC – area under the receiver operating characteristic curve; OR – odds ratio; RR – risk ratio; ICU – intensive care unit; LTC(F) – long-term care (facility); QUADAS – Quality Assessment of Diagnostic Accuracy Studies; RoB – Risk of bias; PLR/NLR – positive/negative likelihood ratio; PPV/NPV – positive/negative predictive value; NS – not stated; efficacy – percentage of correctly classified patients; SENS – sensitivity; SPEC – specificity; SE – standard error; PI – pressure injury; HSROC – hierarchical summary receiver operating characteristic curve; AHCPR – Agency for Health Care Policy and Research; EPUAP – European Pressure Ulcer Advisory Panel; NPUAP – National Pressure Ulcer Advisory Panel; PPPU – Panel for the Prediction and Prevention of Pressure Ulcers; TDCPS – Torrance Developmental Classification of Pressure Sore; RCT – randomised controlled trial; CVD – cardiovascular disease.

[A] or individual study results, where no median was calculated.

#### Table A5. Individual accuracy results of PI risk prediction tools (for which no meta-analysis was conducted*)

| **Tool** (develop-ment year) | **Reviews' authors** (publication year) | **n = no. of studies; N = no. of part-icipants** | **Brief description of included studies** | **Brief description of included study quality** | **Sensitivity (95% CI)** | **Specificity (95% CI)** | **Likelihood ratios (95% CI)** | **AUROC (95% CI)** | **Predictive values (95% CI)** | **Other outcomes** |
|---|---|---|---|---|---|---|---|---|---|---|
| **Andersen**[125] (1982) | Garcia-Fernandez [88] (2014); Pancorbo-Hidalgo[110] (2006) | n = 1;<br>N = 3,398 | Hospital acute ward; classification system not reported but minimum PI stage I; cut-off ≥2; follow-up period 10 days; mean age NS. [110] | Passed CASP quality assessment in both reviews. No further details. | **0.88** [110] | **0.87** [110] | | | **PPV 0.07**<br>**NPV 1.00** [110] | RR = 42.35 (95% CI: 16.67, 107.56) [88]<br><br>'Efficacy' = 83.8%<br>OR = 36.07 (95% CI: 14.07, 92.45) [110] |
| **Arnell**[126] (1983) | Garcia Fernandez[88] (2014) | n = 1;<br>N = 187 | NS | Passed CASP quality assessment. No further details. | | | | | | RR = 4.34 (95% CI: 2.18, 8.63) |
| Braden – modified by Halfens, "**4-factor model**"[127] (2000) | Zhang[108] (2021); Zimmerman[109] (2018); Garcia-Fernandez[88] (2014); Tayyib[104] (2013) | n = 1;<br>N = 53 | Germany; prospective; ICU setting (cardiac surgery); 41.5% female; average age 62 +/- 12.1y; 26 events; EPUAP PI classification used; cut-off ≥2 used. | Passed CASP quality assessment. No further details. [88] | **0.82** [108]<br>**0.85** [104 109] | **0.31** | **PLR 1.19**<br>**NLR 0.58** [108] | | **PPV 0.7**<br>**NPV 0.38** [109] | RR = 1.44 (95% CI: 0.75, 2.75) [88]<br><br>Lower validity for cardiac SICU than Braden [104]. |
| Braden – modified by Halfens, "**extended Braden**"[127] (2000) | Chou[87] (2013) | n = 1;<br>N = 320 | Prospective cohort study; hospital in-patients; no PI on admission; mean age 61y; cut-offs ≤15, 18 used; 'Pressure sore incidence' used as reference standard. | Criteria based on QUADAS-II: Rated as fair quality. The study did not report that the groups received comparable interventions and cut-offs were not pre-defined. | *Cut-off ≤15:*<br>**0.07**<br><br>*Cut-off ≤18:*<br>**0.24** | *Cut-off ≤15:*<br>**0.99**<br><br>*Cut-off ≤18:*<br>**0.95** | *Cut-off ≤15:*<br>**PLR 1.21**<br>**NLR 0.16**<br><br>*Cut-off ≤18:*<br>**PLR 0.83**<br>**NLR 0.14** | | *Cut-off ≤15:*<br>**PPV 0.55**<br>**NPV 0.86**<br><br>*Cut-off ≤18:*<br>**PPV 0.45**<br>**NPV 0.88** | |
| Braden – modified by **Kwong**[115] (2005) | Chou[87] (2013) | n = 2;<br>N = 626 | Two prospective cohort studies; hospital in-patients; no PI on admission; mean ages 58, 79; cut-off points ≤16, ≤19 used; reference standard NPUAP used. | Criteria based on QUADAS-II: Rated good (n=1) and fair quality (n=1; did not report that groups received comparable interventions, blinding of reference standard unclear). Cut-offs were not predefined in either study. | Cut-off ≤16 (n=1):<br>**0.89**<br><br>Cut-off ≤19 (n=1):<br>**0.89** | Cut-off ≤16:<br>**0.75**<br><br>Cut-off ≤19:<br>**0.62** | Cut-off ≤16:<br>**PLR 0.07**<br>**NLR 0.001**<br><br>Cut-off ≤19:<br>**PLR 0.23**<br>**NLR 0.02** | Cut-off ≤19:<br>**0.74 (0.63, 0.84)** | Cut-off ≤16:<br>**PPV 0.07**<br>**NPV 1.0**<br><br>Cut-off ≤19:<br>**PPV 0.19**<br>**NPV 0.98** | |

| Tool (development year) | Reviews' authors (publication year) | n = no. of studies; N = no. of participants | Brief description of included studies | Brief description of included study quality | Sensitivity (95% CI) | Specificity (95% CI) | Likelihood ratios (95% CI) | AUROC (95% CI) | Predictive values (95% CI) | Other outcomes |
|---|---|---|---|---|---|---|---|---|---|---|
| Cubbin & Jackson – revised, "**Jackson & Cubbin**"[117] (1999) | Chou[87] (2013) | n = 3;<br>N = 865 | Three prospective cohort studies; hospital in-patients; mean age ranged between 58 and 62; cut-offs ≤29, 28, 24 used; Sample sizes range between 112 and 534. Reference standards included NPUAP staging system, AHRQ 4-stage criteria and the Stirling Pressure Sore Severity Scale. | Criteria based on QUADAS-II: Rated fair (n=2; sampling method unclear) and good (n=1) quality.<br><br>Blinding of reference standard unclear in all three studies. | *Cut-off ≤29:*<br>**0.83**<br><br>*Cut-off ≤28:*<br>**0.95**<br><br>*Cut-off ≤24:*<br>**0.89** | *Cut-off ≤29:*<br>**0.42**<br><br>*Cut-off ≤28:*<br>**0.82**<br><br>*Cut-off ≤24:*<br>**0.61** | *Cut-off ≤29:*<br>**PLR 0.08**<br>**NLR 0.02**<br><br>*Cut-off ≤28:*<br>**PLR 1.15**<br>**NLR 0.01**<br><br>*Cut-off ≤24:*<br>**PLR 1.03**<br>**NLR 0.08** | *Cut-off ≤29:*<br>**0.72**<br><br>*Cut-off ≤28:*<br>**0.90**<br><br>*Cut-off ≤24:*<br>**0.83** | *Cut-off ≤29:*<br>**PPV 0.07**<br>**NPV 0.98**<br><br>*Cut-off ≤28:*<br>**PPV 0.56**<br>**NPV 0.99**<br><br>*Cut-off ≤24:*<br>**PPV 0.51**<br>**NPV 0.92** | |
| | Tayyib[104] (2013) | n = 3;<br>N = 519 | ICU settings; two prospective observational studies, one longitudinal study; cut-off points ≤29, 28, 24 used. | Not done | *Cut-off ≤29:*<br>NS<br><br>*Cut-off ≤28:*<br>**0.95**<br><br>*Cut-off ≤24:*<br>**0.89** | *Cut-off ≤29:*<br>NS<br><br>*Cut-off ≤28:*<br>**0.82**<br><br>*Cut-off ≤24:*<br>**0.61** | | | | Jackson/Cubbin scale is found to have the highest predictive ability/validity than comparators, in all 3 studies. Comparators: Waterlow, Braden, Douglas, Song and Choi scales. |
| **COMHON**[128] (2011) | Zhang[108] (2021) | n = 1;<br>N = 2,777 | Spain; retrospective; ICU setting; 38.1% female; average age 63 +/- 16y; 154 events; NPUAP PI classification used; cut-off >12 used. | Reported overall, not by scale. | **0.83** | **0.52** | **PLR 1.72**<br>**NLR 0.33** | **0.70** | | |
| **Compton**[129] (2008) | Garcia-Fernandez[88] (2014) | n = 1;<br>N = 698 | NS | Passed CASP quality assessment. No further details. | | | | | | RR = 4.85 (95% CI: 3.66, 6.42) |
| **Douglas**[130] (1986) | Zhang[108] (2021); Zimmerman[109] | n = 1;<br>N = 112 | Korea; prospective, longitudinal; ICU (surgical, internal or neurological) | Passed quality assessment in two reviews that utilised CASP[88 110]. | **1.00** | **0.18** | **PLR 1.22** [A]<br>**NLR 0.00** | **0.79** | **PPV 0.34**<br>**NPV 1.00** | RR = 10.76 (95% CI: 0.70, 166.27) [88] |

| Tool (develop-ment year) | Reviews' authors (publication year) | n = no. of studies; N = no. of part-icipants | Brief description of included studies | Brief description of included study quality | Sensitivity (95% CI) | Specificity (95% CI) | Likelihood ratios (95% CI) | AUROC (95% CI) | Predictive values (95% CI) | Other outcomes |
|---|---|---|---|---|---|---|---|---|---|---|
| | (2018); Garcia-Fernandez[88] (2014); Tayyib[104] (2013); Chou[87] (2013); Pancorbo-Hidalgo[110] (2006) | | setting; history of PIs unclear; 32.8% female; mean age 62y; 35 events, with follow-up to discharge/moved to other ward/death[110]; 'Panel for the Prediction and Prevention of Pressure Ulcers' classification used[108]; NPUAP classification used[87] (minimum stage I); cut-off ≤18 used. | Criteria based on QUADAS-II: Rated as good quality. Blinding of reference standard unclear[87]. | | | | | | 'Efficacy' = 43.6%[110]<br><br>Lower validity than Jackson/Cubbin[104]. |
| **DUPA**[131] (1995) | Garcia-Fernandez[88] (2014) | n = 1; N = 85 | NS | Passed CASP quality assessment. No further details. | | | | | | RR = 2.13 (95% CI: 1.21, 3.75) |
| **Dutch CBO**[132] (1992) | Chou[87] (2013) | n = 1; N = 220 | Retrospective study in a nursing home. Mean age 79. Dutch CBO cut-off of ≤10 used. NPUAP staging system used as reference standard. | Criteria based on QUADAS-II: Rated as fair quality. Sampling method unclear, study did not report that interventions were comparable, the scale cut-offs were not pre-defined, it was unclear whether the reference standard was applied to all patients, whether the same reference standard was applied to all patients and whether it was blinded. | 0.55 | 0.75 | | | | |
| **EMINA**[118] (2001) | Pancorbo-Hidalgo[110] (2006) | n = 1; N = 673 | Hospital; classification system not reported but minimum stage I; EMINA cut-off ≥4; follow-up period 7 days; average age NS. | All studies considered to be 'valid' through use of CASP. | **0.77** | **0.72** | | **0.82** | **PPV 0.17**<br>**NPV 0.98** | 'Efficacy' = 71.9%;<br>OR = 8.24 (95% CI: 4.10, 16.54) |
| | Zhang[108] (2021) | n = 1; N = 189 | Spain; prospective; ICU setting; 32.8% female; average age 59.4 +/- 16.9y; | Reported overall, not by scale. | **0.94** | **0.33** | **PLR 1.41**<br>**NLR 0.17** | **0.64** | | |

| Tool (develop-ment year) | Reviews' authors (publication year) | n = no. of studies; N = no. of part-icipants | Brief description of included studies | Brief description of included study quality | Sensitivity (95% CI) | Specificity (95% CI) | Likelihood ratios (95% CI) | AUROC (95% CI) | Predictive values (95% CI) | Other outcomes |
|---|---|---|---|---|---|---|---|---|---|---|
| | | | 53 events; NPUAP-EPUAP PI classification used; cut-off point of >10 | | | | | | | |
| **Fragm-ment**[133] (2002) | Garcia-Fernandez[88] (2014); Chou[87] (2013); Pancorbo-Hidalgo[110] (2006) | n = 1; N = 1,190 | Prospective cohort study; hospital in-patients (medical units, surgical units and ICU); no PIs on admission; mean age 61y; cut-off >3 used; NPUAP staging system used as reference standard (minimum stage I); follow-up period 3 weeks/to discharge. | Passed quality assessment in two reviews that utilised CASP[88 110].<br><br>Criteria based on QUADAS-II: Rated as fair quality, but scale was evaluated on the same population as it was developed on, authors did not report that groups received comparable interventions, cut-offs not pre-defined and blinding of reference standard unclear[87]. | **0.62** | **0.85** | **PLR 0.73**<br>**NLR 0.08** | **0.79 (0.75, 0.82)** | **PPV 0.34**<br>**NPV 0.95** | RR = 5.74 (95% CI: 4.40, 7.50) [88]<br><br>RR = 1.6 (95% CI: 1.4, 1.7) per 1 point increase in score[87]. |
| **Gosnell**[134] (1973) | Zhang[108] (2021); Garcia-Fernandez[88] (2014); Chou[87] (2013) | n=1; 230 | Iran; prospective; ICU in-patients; no PIs on admission; 56.5% female; average age 60y; 74 events; AHCPR PI classification used; cut-off point of 16 | Passed CASP quality assessment[88].<br><br>Criteria based on QUADAS-II: Rated as fair quality. Sampling method and blinding of reference standard unclear[87]. | **0.85** | **0.83** | **PLR 4.92**<br>**NLR 0.18** | | | |
| **Hatanaka** [**135**] (2008) | Chou[87] (2013) | n = 1; N = 149 | Prospective cohort study; hospital in-patients; no PIs on admission; mean age 72y; cut-off 0.28 (possible range 0-1) used; own criteria used for reference standard. | Criteria based on QUADAS-II: Rated as fair quality. The scale was evaluated in the same population as it was developed on, sampling method unclear, the scale cut-offs were not pre-defined, unclear whether the reference standard was | **0.73** | **0.70** | **PLR 0.85**<br>**NLR 0.14** | **0.79** | **PPV 0.46**<br>**NPV 0.88** | |

| **Tool** (develop-ment year) | **Reviews' authors** (publication year) | **n = no. of studies; N = no. of part-icipants** | **Brief description of included studies** | **Brief description of included study quality** | **Sensitivity (95% CI)** | **Specificity (95% CI)** | **Likelihood ratios (95% CI)** | **AUROC (95% CI)** | **Predictive values (95% CI)** | **Other outcomes** |
|---|---|---|---|---|---|---|---|---|---|---|
| | | | | credible and whether it was blinded. | | | | | | |
| **HPUR**[136] (2003) | Garcia-Fernandez[88] (2014) | n = 1; N = 54 | NS | Passed CASP quality assessment. No further details. | | | | | | RR = 3.02 (95% CI: 1.26, 7.20) |
| **Knoll**[137] (1988) | Garcia-Fernandez [88] (2014); Chou[87] (2013); Pancorbo-Hidalgo[110] (2006) | n = 1; N = 60 | Prospective cohort study; LTCF; no PIs on admission; mean age 81y; cut-off 12; reference standard unclear, but minimum stage I; follow-up period 28 days. | Passed quality assessment in two reviews that utilised CASP[88 110].<br><br>Criteria based on QUADAS-II: Rated as fair quality. Unclear whether the scale was evaluated in the same population as it was developed on, unclear whether the reference standard was credible and whether reference standard was blinded[87]. | **0.86** | **0.56** | **PLR 1.71**<br>**NLR 0.22** | | **PPV 0.63**<br>**NPV 0.82** | RR = 3.47 (95% CI: 1.39, 8.71) [88]<br><br>'Efficacy' = 70.0% OR = 7.71 (95% CI: 2.17, 27.42)[110] |
| Norton – modified by **Bale**[138] (1995) | Garcia-Fernandez [88] (2014) | n = 1; N = 240 | NS | Passed CASP quality assessment. No further details. | | | | | | RR = 1.35 (95% CI: 0.68, 2.71) |
| | Chou[87] (2013) | n = 1; N = 79 | Prospective cohort study on hospice patients. The scale was reversed so that a higher score represents higher risk. Mean age was 67y. Modified Norton cut-off of >10 used. Torrance Developmental Classification of Pressure Sores used as reference standard. | Criteria based on QUADAS-II: Rated as fair quality. The study didn't report that the groups received comparable interventions, the cut-offs were not pre-defined and blinding of reference standard unclear. | **1.00** | **0.31** | **PLR 3.20**<br>**NLR 0.00** | | **PPV 0.04**<br>**NPV 1.00** | |
| Norton – modified by **Bienstein**[122] (1991) | Zhang[108] (2021); Tayyib[104] | n = 1; N = 53 | Germany; prospective; ICU setting (cardiac surgery); 41.5% female; average age 62 +/- 12.1y; 26 events; | Criteria based on QUADAS-II: Rated as fair quality. The study didn't report that the group received comparable | *Cut-off ≤21: 0.33*<br><br>*Cut-off ≤23* | *Cut-off ≤21: 0.94*<br><br>*Cut-off ≤23* | *Cut-off ≤21: PLR 0.92 NLR 0.68* | | *Cut-off ≤21: PPV 0.92 NPV 0.40* | Lower validity for cardiac SICU than Braden[104]. |

| **Tool** (develop-ment year) | **Reviews' authors** (publication year) | **n = no. of studies; N = no. of part-icipants** | **Brief description of included studies** | **Brief description of included study quality** | **Sensitivity (95% CI)** | **Specificity (95% CI)** | **Likelihood ratios (95% CI)** | **AUROC (95% CI)** | **Predictive values (95% CI)** | **Other outcomes** |
|---|---|---|---|---|---|---|---|---|---|---|
| | (2013); Chou[87] (2013) | | EPUAP PI classification used; cut-off points ≤25 (and ≤21, 23[87]). | interventions and the cut-offs were not pre-defined[87]. | *0.41*<br><br>*Cut-off ≤25:* 0.58 | *0.88*<br><br>*Cut-off ≤25:* 0.47 | *Cut-off ≤23: PLR 0.88 NLR 0.64*<br><br>*Cut-off ≤25:* PLR 1.11 NLR 0.87 | | *Cut-off ≤23: PPV 0.88 NPV 0.42*<br><br>*Cut-off ≤25:* PPV 0.70 NPV 0.35 | |
| Norton – modified by **Stotts**[139] (1988) | Chou[87] (2013) | n = 1;<br>N = 387 | Prospective cohort study on surgical in-patients. Mean age was 53. Modified Norton cut-off of ≤14. Used own criteria for reference standard. | Criteria based on QUADAS-II: Rated as fair quality. The study didn't report that the groups received comparable interventions, cut-offs were not pre-defined, sampling method unclear and blinding of reference standard interpretation unclear. | **0.16** | **0.95** | **PLR 0.67**<br>**NLR 0.18** | | **PPV 0.4**<br>**NPV 0.85** | |
| Norton – modified by **Ek**[140] (1997) | Pancorbo-Hidalgo[110] (2006) | n = 1;<br>N = 81 | Hospital orthopaedics; classification system not reported but minimum stage I; cut-off ≤21 used; follow-up 14 days after surgery/to discharge; average age 82y. | All studies considered to be 'valid' through use of CASP. | **0.71** | **0.44** | | | **PPV 0.35**<br>**NPV 0.78** | 'Efficacy' = 52.0% OR = 1.92 (95% CI: 0.69, 5.36) |
| **NOVA-4**[141] (1999) | Garcia-Fernandez[88] (2014) | n = 1;<br>N = 187 | NS | Passed CASP quality assessment. No further details. | | | | | | RR = 4.72 (95% CI: 1.90, 11.77) |
| **PSPS**[123] (1987) | Pancorbo-Hidalgo[110] (2006) | n = 1;<br>N = 1,244 | Hospital orthopaedics; classification system and minimum PI stage not reported; PSPS cut-off >6; follow-up period 3 weeks; average age NS. | All studies considered to be 'valid' through use of CASP. | **0.89** | **0.76** | | | **PPV 0.14**<br>**NPV 0.99** | 'Efficacy' = 76.6% OR = 25.62 (95% CI: 10.73, 61.20) |
| **RAPS**[142] (2002) | Zhang[108] (2021); Zimmerman[109] (2018) | n = 1;<br>N = 122 | Turkey; prospective; ICU setting; 57.4% female; average age 56.5 +/- 18.6y; 31 events; NPUAP PI | Reported overall, not by scale. | **0.74** | **0.32** | **PLR 1.09**<br>**NLR 0.81** | **0.5** | **PPV 0.387**<br>**NPV 0.913** | Studies that compared the predictive capacity of |

| Tool (development year) | Reviews' authors (publication year) | n = no. of studies; N = no. of participants | Brief description of included studies | Brief description of included study quality | Sensitivity (95% CI) | Specificity (95% CI) | Likelihood ratios (95% CI) | AUROC (95% CI) | Predictive values (95% CI) | Other outcomes |
|---|---|---|---|---|---|---|---|---|---|---|
| | | | classification used; cut-off point of ≥27 | | | | | | | specific and generic scales (according to setting) presented significant variation. Sensitivity variation of all scales was high, ranging from 60% to 100%. However, in specificity values, the ICU specific scales showed higher variations (from 61% to 86%), when compared with generic scales (from 5% to 69%)[109]. |
| | Garcia-Fernandez[88] (2014); Chou[87] (2013); Pancorbo-Hidalgo[110] (2006) | n = 1; N = 488 | Prospective cohort; hospital in-patients (hospital medical, surgical, orthopaedic and geriatric units); no PIs on admission; mean age 70y; cut-off ≤36; own criteria used for reference standard; minimum stage I; follow-up period 12 weeks. | Passed quality assessment in two reviews that utilised CASP[88 110].<br><br>Criteria based on QUADAS-II: Rated as poor quality. The scale was evaluated in the same population as it was developed on, the test cut-offs were not pre-specified, the study did not report that groups received comparable interventions and attrition | **0.57** | **0.58** | **PLR 0.19**<br>**NLR 0.10** | | **PPV 0.14**<br>**NPV 0.92** | RR = 1.71 (95% CI: 1.03, 2.85) [88]<br><br>'Efficacy' = 57.4% OR = 1.82 (95% CI: 1.06, 3.11)[110] |

| Tool (develop-ment year) | Reviews' authors (publication year) | n = no. of studies; N = no. of part-icipants | Brief description of included studies | Brief description of included study quality | Sensitivity (95% CI) | Specificity (95% CI) | Likelihood ratios (95% CI) | AUROC (95% CI) | Predictive values (95% CI) | Other outcomes |
|---|---|---|---|---|---|---|---|---|---|---|
| | | | | was high. Sampling method and whether the reference standard was blinded were unclear. | | | | | | |
| **SS scale**[143] (2008) | Zhang[108] (2021); Zimmerman[109] (2018); Garcia-Fernandez[88] (2014); Tayyib[104] (2013) | n = 1; N = 253 | Indonesia; prospective; ICU setting; 37.5% female; 72 events; NPUAP PI classification used; cut-off point >4 | Passed CASP quality assessment. No further details.[88] | **0.81** | **0.83** | **PLR 4.70** **NLR 0.23** | **0.89** | **PPV 0.65** **NPV 0.91** | RR = 7.63 (95% CI: 4.52, 12.89) [88] |
| **Sunderland** [144] (1995) | Zhang[108] (2021); Zimmerman[109] (2018) | n=1; 90 | Portugal; prospective; ICU setting; 36.7% female; average age 70y; 15 events; NPUAP-EPUAP PI classification used; cut-off point of 28 | Reported overall, not by scale. | **0.6** | **0.87** | **PLR 4.50** **NLR 0.46** | **0.86** | **PPV 0.47** **NPV 0.92** | |
| | Garcia-Fernandez[88] (2014) | n = 1; N = 15 | NS | Passed CASP quality assessment. No further details. | | | | | | RR = 1.25 (95% CI: 0.22, 7.08) |
| **TNH-PUPP**[145] (2011) | Chou[87] (2013) | n = 1; N = 165 | Prospective cohort study in hospital general wards, critical care or emergency departments. History of PI unclear; existing PI (%) unclear. Mean age 68y. TNH-PUPP cut-off 3. Reference standard unclear. | Criteria based on QUADAS-II: Rated as fair quality. Sampling method unclear, the study did not report that interventions were comparable, the scale cut-offs were not pre-defined, it was unclear whether the reference standard was credible and whether it was blinded. | **0.86** | **0.73** | **PLR 0.13** **NLR 0.01** | **0.90 (0.82, 0.99)** | **PPV 0.13** **NPV 0.99** | |
| **Watkinson** [146] (1997) | Garcia-Fernandez[88] (2014) | n = 1; N = 185 | NS | Passed CASP quality assessment. No further details. | | | | | | RR = 34.88 (95% CI: 2.12, 574.39) |

| Tool (develop-ment year) | Reviews' authors (publication year) | n = no. of studies; N = no. of part-icipants | Brief description of included studies | Brief description of included study quality | Sensitivity (95% CI) | Specificity (95% CI) | Likelihood ratios (95% CI) | AUROC (95% CI) | Predictive values (95% CI) | Other outcomes |
|---|---|---|---|---|---|---|---|---|---|---|
| Bayesian Network: **ML Kaewprag [2]**[147] (2017) | Qu[103] (2022) | n = 1; N = 7,717 | NS; Sensitivity and specificity calculated by umbrella review team from raw data | QUADAS-II: Low overall risk of bias. | **0.64 (0.60, 0.68)** n = 590 | **0.81 (0.80, 0.82)** n = 7127 | | | | |
| Bayesian Network: **ML Ladios Martin**[148] (2020) | Qu[103] (2022) | n = 1; N = 1,769 | NS; Sensitivity and specificity calculated by umbrella review team from raw data | QUADAS-II: High overall risk of bias. High risk of bias and applicability concerns in reference standard domain. | **0.04 (0.09, 0.12)** n = 68 | **0.93 (0.92, 0.94)** n = 1701 | | | | |
| **LOS** model: **ML Kim [2]**[149] (2006) | Qu[103] (2022) | n = 1; N = 2,347 | NS; Sensitivity and specificity calculated by umbrella review team from raw data | QUADAS-II: Low overall risk of bias. | **0.92 (0.84, 0.97)** n = 84 | **0.67 (0.65, 0.69)** n = 2263 | | | | |

CI – confidence interval; AUROC – area under the receiver operating characteristic curve; NS – not stated, CASP – Critical Appraisal Skills Programme; RR – risk ratio; efficacy – percentage of correctly classified patients; OR – odds ratio; PI – pressure injury; (S)ICU – (surgical) intensive care unit; QUADAS – Quality Assessment of Diagnostic Accuracy Studies; EPUAP – European Pressure Ulcer Advisory Panel; NPUAP – National Pressure Ulcer Advisory Panel; AHCPR – Agency for Health Care Policy and Research; AHRQ – Agency for Healthcare Research and Quality; PLR – positive likelihood ratio; NLR – negative likelihood ratio; PPV – positive predictive value; NPV – negative predictive value; LTC(F) – long-term care (facility); ML – machine learning; LOS – abbrev. not defined within review.

[A] re-calculated due to discrepancy between reviews. * Three reviews [87 88 98] conducted inappropriate statistical syntheses, therefore for tools included in those syntheses, the results of other individual studies of such tools may also be presented within this table.

#### Table A6. Results from all 11 included systematic reviews evaluating clinical effectiveness

| Review author (publication year) | Tools included | Setting of included studies; study design; sample size | Included outcomes | Brief description of study quality | Relevant results from included studies |
|---|---|---|---|---|---|
| Lovegrove[94] (2021) | Braden; Maelor score; Norton; Ramstadius; Waterlow | Acute care hospital n=1, inpatient units n=1, ICU n=1, internal medicine and oncology wards n=1;<br><br>Design: cross-sectional survey n=2, RCT n=1, observational inter-rater reliability n=1;<br><br>Sample size 45 to 1231 | PI risk scores; PI incidence; PI preventative interventions; interrater reliability | RoB assessed using JBI tools or analytical cross-sectional study appraisal checklist. The RCT was judged as high quality. Of the remaining studies, two were judged as high quality and one as moderate quality; inclusion criteria not clearly stated and no strategies to deal with confounding. | • The Braden scale had the highest ICC across two ICUs (0.72 and 0.84), followed by subjective assessments (0.51 and 0.71) then the Waterlow score (0.36 and 0.51) (Kottner and Dassen 2010).<br>• There were no differences in patient management ('pressure care plan' and use of a special mattress) based on PI risk assessment method (clinical judgement, Ramstadius tool or Waterlow score). PI incidence difference between groups not significant (p=0.44) (Webster 2011).<br>• A hospital that used the Maelor scale reported a higher rate of PI preventative strategies than a site that used nurses' clinical judgement (Moore 2015).<br>• 33% of nursing assessments differed from those of the computer-generated Norton score. Of the patients assessed as being at high risk of PI by the Norton scale, 64% were repositioned 2 or 3 hourly, 7% were repositioned 4 or 5 hourly, 4% were repositioned hourly and 7% were not repositioned at all (Voz 2011). |
| Mehicic[96] (2024) | Braden | All studies were based in ICU settings;<br><br>Design: all prospective studies n=9;<br><br>Sample size 3 to 4137 | Internal consistency (n=2); interrater reliability (n=6); measurement error (n=3); convergent validity (n=3) | RoB assessed using COSMIN RoB checklist.<br><br>All studies assessing internal consistency or measurement error scored 'Very Good' for all relevant criteria.<br><br>Half (n=3) the studies assessing interrater reliability scored 'Very Good' for all relevant criteria; one study scored 'Doubtful' for all relevant criteria, one study scored 'Inadequate' for 4/7 criteria, and the last study scored 'Doubtful' for 2/5 criteria and 'Inadequate' for 1/5 criteria. | • **Internal consistency.** Two studies (Lima-Serrano 2018 and Adibelli 2019) reported Cronbach's alpha ranging from poor (0.43) to good (0.85).<br>• **Interrater reliability**. Only four (4/6) studies reported intraclass correlation, ranging from 0.66 to 0.96 for Braden sum score. One study (Veiga 2022) reported weighted kappa = 0.17 and another reported kappa ranging from 0 to 0.86 (not weighted) (Simao 2013).<br>• **Measurement error.** Standard error of measurement (SEM) was around 2 points of the sum score (1.83 and 1.67–1.64) in two studies (Fullbrook 2016 and Kottner 2010, respectively), and a slightly smaller SEM of 1.31 was reported by the third study (Veiga 2022).<br>• **Convergent validity.** The Braden scale was compared with four comparator tools: the Cubbin-Jackson scale (Delawder 2021), the COHMON Index and Norton scale (Fulbrook 2016), and the Waterlow score (Fulbrook 2016; Kottner 2010).<br>• One study found weak correlation between the sum scores of the Braden scale and Waterlow scale (r=0.22) (Fullbrook 2016), whereas another study found strong correlations between the two scales (r=0.72,0.71) (Kottner 2010). |

| **Review author** (publication year) | **Tools included** | **Setting of included studies; study design; sample size** | **Included outcomes** | **Brief description of study quality** | **Relevant results from included studies** |
|---|---|---|---|---|---|
| | | | | Two studies assessing convergent validity scored 'Very Good' for all criteria; the other study scored 'Doubtful' for 3/4 criteria. | • Fullbrook 2016 reported strong correlations between the Braden and Norton sum scores (r=0.77), and between the Braden and the COMHON Index (r=0.70).<br>• Delawder 2021 reported strong correlation between the Braden and the Cubbin-Jackson scale (r=0.80) |
| Moore[97] (2019) | Braden; Waterlow; Ramstadius | Military hospital n=1, internal medicine and oncology wards n=1;<br><br>Design: RCT n=1, cluster randomised trial n=1;<br><br>Sample sizes 286 and 1231 | PI incidence; severity of PIs | RoB assessed using Cochrane tool (Higgins (2011)).<br>Both studies at high RoB due to blinding issues. One study at RoB also due to baseline imbalance and incorrect analyses. | • No differences in PI incidence when using Braden scale or clinical judgement (Braden vs. clinical judgement+training, RR 0.97, 95% CI 0.53, 1.77; Braden vs clinical judgement RR 1.43, 95% CI 0.77, 2.68) (Saleh 2009).<br>• No difference in PI incidence when using a risk assessment tool compared to clinical judgement (RR 1.10, 95% CI 0.68, 1.81 and RR 0.79, 95% CI 0.46, 1.35, for Waterlow and Ramstadius respectively) (Webster 2011).<br>• No difference in PI severity based on risk assessment tools vs. clinical judgement (Webster 2011). |
| Gaspar[89] (2019) | Waterlow; Ramstadius | Internal medical and oncology wards n=1;<br><br>Design: RCT n=1<br><br>Sample size 1231 | PI incidence | RoB assessed using Evidence-Based Librarianship Critical Appraisal checklist. Overall validity of the eligible study was 95.83%. The study scored 87.5% for the population domain. | • The incidence of HAPI was similar between the groups (6.8% vs. 7.5% vs. 5.4%, p=0.44 for Waterlow, Ramstadius and clinical judgement respectively) (Webster 2011). |
| Chou[87] (2013) | Norton modified by Bale; Braden; Waterlow; Ramstadius | Hospital n=2, hospice n=1;<br><br>Design: non-randomised n=1, cluster randomised trial n=1, RCT n=1;<br><br>Sample size 240 to 1231 | PI incidence, severity of PIs; PI preventative interventions | RoB assessed with criteria consistent with AHRQ Methods Guide for Effectiveness and Comparative Effectiveness Reviews. One RCT was rated as good quality and the other as poor due to randomisation and blinding issues. The cohort study was rated as poor; there were blinding issues and confounding was not investigated. | • No difference in PI incidence when using a risk assessment tool compared to clinical judgement (RR 1.10, 95% CI 0.68, 1.81 and RR 0.79, 95% CI 0.46, 1.35, for Waterlow and Ramstadius respectively) (Webster 2011).<br>• The modified version of the Norton scale with use of preventive interventions is associated with lower risk of PIs compared with clinical judgment (RR 0.11, 95% CI 0.03, 0.46) (Bale 1995).<br>• No difference in risk of PIs when one of three interventions was used (22% vs. 22% vs. 15%, p=0.38 for nurse training+mandatory Braden scale, nurse training+optional Braden scale and no training respectively) (Saleh 2009). |
| Pancorbo-Hidalgo[110] (2005) | Norton; Norton modified by | Hip fracture inpatients n=1, palliative care/hospice n=1, neurosurgery, general | PI incidence; PI preventative interventions | RoB assessed using CASP Guide for clinical trials or | • Compared a strategy that gave high risk patients (based on modified Norton score) a risk alarm sticker to standard care. No significant |

| **Review author** (publication year) | **Tools included** | **Setting of included studies; study design; sample size** | **Included outcomes** | **Brief description of study quality** | **Relevant results from included studies** |
|---|---|---|---|---|---|
| | Bale; Norton modified by Ek 97 | medicine, orthopaedic, and oncology units n=1;<br><br>Design: prospective controlled (contemporaneous controls) n=1, before-and-after n=1;<br><br>Sample size: 124 to 223 | | the critical assessment guide developed for the clinical practice guide for PI assessment and prevention for cohort studies (Rycroft-Malone & McInness (2000)). Studies excluded if considered not to be valid. | difference between the groups in the incidence of PIs (OR 1.18, 95% CI 0.52, 2.66) (Gunningberg 1999).<br>• The modified version of the Norton scale with use of pressure-reducing mattresses is associated with lower risk of PIs compared with clinical judgment (decrease in PI incidence in the Norton scale group of 19.8%, 95% CI 12.2, 27.4). More patients in the Norton group were given then mattresses (Bale 1995).<br>• Compared the Norton scale with training to standard care. There was a significant difference in the number of preventative interventions (18.96 vs. 10.75, for Norton and usual care respectively) (Hodge 1990). |
| Health Quality Ontario[91] (2009) | Norton; Norton modified by Bale; Norton modified by Ek 97 | Hip fracture inpatients n=1, palliative care/hospice n=1, neurosurgery, general medicine, orthopaedic, and oncology units n=1;<br><br>Design: prospective controlled (contemporaneous controls) n=1, before-and-after n=1;<br><br>Sample size 124 to 223 | PI incidence; PI preventative interventions | RoB assessment criteria name not given. Two studies met 6/8 and one study met all quality assessment requirements. In the studies that didn't meet all requirements, there were blinding and loss to follow-up issues. One study used a version of the Norton scale that was not validated. | • Compared a strategy that gave high risk patients (based on modified Norton score) a risk alarm sticker to standard care. No significant difference between the groups in the incidence of PIs (Gunningberg 1999).<br>• Compared a strategy where patients received a pressure support system allocated according to the modified Norton scale to one where the nurse chose whether to give a special mattress. Using the scale significantly reduced the incidence of PIs (22.4% vs. 2.5%, p<0.001) (Bale 1995).<br>• Compared the Norton scale with training to standard care. There was a significant difference in the number of preventative interventions (18.96 vs. 10.75, for Norton and usual care respectively). Interventions were used earlier for Norton vs. usual care (on day 1, 61% vs. 50%, p<0.002). No significant difference in the incidence of PIs between the groups (Hodge 1990). |
| Lovegrove[95] (2018) | Braden; Cubbin & Jackson; modified Norton; Ramstadius; Waterlow | Hospital or acute care n=20;<br><br>Design: cross-sectional n=15, observational n=2, RCT n=1, pre-test post-test n=1, prospective cohort=1;<br><br>Sample size not clearly reported | Level of risk in relation to the prescription of PI preventive interventions | RoB assessed using JBI tools. The RCT, eleven cross-sectional studies, the cohort and the observational studies were rated as high quality (n=15). Four cross-sectional and the pre-test post-test studies were rated as moderate quality (n=5). Reporting of the prescription of interventions for PI was unclear. | • Limited mention of specific risk assessment tools.<br>• Seven studies reported PI risk assessment, risk status and interventions implemented for patients identified as being at-risk in some way and 13 linked it to preventative intervention prescription and implementation in some way.<br>• No studies linked PI risk assessment to preventative intervention prescription alone. |

| **Review author** (publication year) | **Tools included** | **Setting of included studies; study design; sample size** | **Included outcomes** | **Brief description of study quality** | **Relevant results from included studies** |
|---|---|---|---|---|---|
| Baris[84] (2015) | Braden | ICU n=12, general hospital n=2, surgical n=1, orthopaedics and traumatology clinic n=1;<br><br>Design: descriptive n=11, experimental n=5;<br><br>Sample size 22 to 422 | PI incidence; reliability of Braden scale | No RoB assessment | • PIs developed in 64% of patients with a Braden score ≤12 (Bakanoglu 2010).<br>• PIs developed in 20.8% of patients with a Braden score ≥13 and in 46.6% of patient with a Braden score ≤12. The difference was statistically significant (Oguz 1998).<br>• Braden scale had a reliability coefficient of 0.95 (Oguz 1998).<br>• Seven studies regarded the Braden scale as 'valid and reliable'. |
| Kottner[93] (2009) | Waterlow; Waterlow 12-item | Hospital n=6, community n=1, unclear n=2;<br><br>Design: interrater reliability n=7 studies, intra-rater reliability n=1, unclear n=1;<br><br>Sample size 1 to 52 | Interrater reliability; intra-rater reliability | Used own RoB assessment criteria. Two studies rated as high quality. Issues with the other studies include poor description of raters, subjects, methods, procedures and results, non-independent scoring, non-representative results and inappropriate statistical methods. | • Waterlow inter-rater reliability: $p^a$ within one point=0.6 (Dealey 1989)<br>• Waterlow inter-rater reliability: $p^a$=0.25; $p^a$ within one point=0.5; range of differences 0-11 (Edwards 1995)<br>• Waterlow inter-rater reliability: $p^a$=0; $p^a$ within one point=0.11; $p^a$ within two points=0.33; $p^a$ within 3 points=0.44; $p^a$ within 4 points=0.56 (Watkinson 1996)<br>• Waterlow inter-rater reliability: r=0.99 (Pang and Wong 1998)<br>• Waterlow 12-item inter-rater reliability: $p^a$=0.21-0.57; $p^a$ within 1 point=0.29-0.72; $p^a$ within 2 points=0.5-0.86; range of differences 0-15 (Cook 1999)<br>• Waterlow inter-rater reliability: mean $p^a$=78.1; range of differences 0-16 (Hale 1999)<br>• Waterlow inter-rater reliability: $p^a$=0.11; $p^a$ within one point=0.27; $p^a$ within 2 points 0.4; range of differences 0-24 (Kelly 2005)<br>• Waterlow intra-rater reliability: $p^a$=55.1; ICC=0.97 (95% CI 0.94, 0.98) (Hale 1999)<br>• Waterlow type of comparison unclear: ICC=0.95 (Jalali and Rezaie 2005) |
| Tayyib[104] (2013) | Braden; Waterlow; SS scale | ICU n=2; ICU cardiac surgery n=1;<br><br>Design: prospective cohorts n=2, observational n=1;<br><br>Sample size 105 to 3027 | Reliability of PI scales; interrater reliability | No RoB assessment | • Braden scale demonstrated high reliability (Pearson's r: 0.83-0.99) (Lewicki 2000).<br>• Braden scale has high inter-rater reliability value compared to Waterlow scale (Kottner and Dassen 2010).<br>• S.S. showed high inter-rater reliability (r = 1) (Suriadi 2007). |

AHRQ – Agency for Healthcare Research; CASP – Critical Appraisal Skills Checklist; COSMIN – COnsensus-based Standards for the selection of health Measurement INstruments; ICU – Intensive Care Unit; JBI – Joanna Briggs Institute; RCT – Randomised Controlled Trial; RoB – Risk of Bias; S.S. – Suriadi Sanada Scale

[A]Proportion of agreement.
